# Explainability in action: A metric-driven assessment of local explanations for healthcare tabular models

**DOI:** 10.1101/2025.05.20.25327976

**Authors:** M. Atif Qureshi, Abdul Aziz Noor, Awais Manzoor, Muhammad Deedahwar Mazhar Qureshi, Arjumand Younus, Wael Rashwan

**Affiliations:** ADAPT Centre, Ireland; Research Ireland Centre for Research Training in Machine Learning (ML-Labs); eXplainable Analytics Group, Faculty of Business, Technological University Dublin; School of Information and Communication Studies, University College Dublin; Maynooth University, School of Business

## Abstract

Explainable AI (XAI) is increasingly used in clinical machine learning, yet quantitative evaluation of explanation quality is often reported inconsistently across methods and datasets. We present a reproducible, metric-driven framework for evaluating XAI methods on healthcare tabular data. The framework consolidates six established, family-specific metrics, fidelity, simplicity, consistency, robustness, precision, and coverage, into explicit equations; pairs them with a pre-specified focal-model protocol; and releases open-source code with a method–metric applicability map. We evaluate LIME, SHAP, Anchors, EBM, and TabNet across four public healthcare tabular datasets. Post-hoc explainers are applied to a single selected Random Forest focal predictor to control model-induced variability, whereas EBM and TabNet are assessed through their native interpretability mechanisms. Global explanation summaries are reported descriptively only. The results show that SHAP/TreeSHAP provides exact score reconstruction for the Random Forest setting, while LIME produces simpler but lower-fidelity explanations with greater instance-level variability. LIME and SHAP show the strongest rank agreement among the evaluated pairs, although agreement varies across datasets. TabNet often yields compact native explanations, but these must be interpreted alongside its dataset-specific predictive performance. EBM and TabNet show low sensitivity under the fixed Gaussian-jitter robustness protocol, while Anchors produces high-precision rules with reduced coverage at stricter thresholds. Overall, the framework enables controlled comparison under explicit method–metric and focal-model assumptions, supporting more transparent XAI selection for tabular machine learning. Although demonstrated in healthcare, the framework is transferable to other high-stakes tabular domains. Source code: https://github.com/matifq/XAI_Tab_Health.

## Introduction

Machine learning (ML) continues to play a growing role in healthcare, supporting tasks such as diagnosis, prognosis, and treatment planning. However, the lack of transparency in many ML models remains a major barrier to clinical adoption [1]. In high-stakes domains like healthcare, explainability is essential not only for regulatory compliance and ethical deployment, but also for building trust among clinicians and patients [2].

Healthcare data are often tabular in nature, particularly in applications involving electronic health records (EHRs), clinical registries, and structured survey data. Despite the prevalence of tabular data in real-world clinical systems, much of the XAI literature has focused on image- and text-based modalities. While many studies advocate the use of XAI methods, comparatively few provide systematic empirical comparisons, and even fewer evaluate multiple explainers using clearly defined and practically meaningful quantitative metrics [1]. This gap limits the ability of practitioners and data scientists to make informed decisions about which XAI methods are appropriate for their specific use cases.

This article addresses this gap by benchmarking five widely used XAI methods, SHAP, LIME, Anchors, EBM, and TabNet, on four healthcare tabular datasets. The study is methodological in scope and does not aim to derive, validate, or recommend clinical decision rules; instead, it focuses on standardized quantitative evaluation of explainability methods. Based on formal definitions from the XAI literature, we operationalize and implement six established evaluation metrics: fidelity, simplicity, consistency, robustness, precision, and coverage. These metrics are applied systematically across methods and datasets to enable controlled comparison.

### Study objective and scope

This study focuses on the methodological evaluation of explainable AI techniques for tabular healthcare models, rather than on addressing a specific clinical decision task or proposing new explainability methods. Specifically, it addresses a methodological gap in the XAI literature: the lack of a standardized, reproducible protocol for quantitatively evaluating and comparing explainability methods on tabular healthcare data. The novelty of the contribution lies at the framework level, through standardized operationalization of established metrics, explicit mapping of metrics to explainer families, control of model-induced confounds via a pre-specified focal-model protocol, and consistent application across multiple healthcare datasets with released code. The focal-model protocol is used to control model-induced variability in post-hoc evaluation; the study does not aim to characterize explainer–model sensitivity across all candidate predictors, which is treated as a future extension. This study is explicitly positioned as a methodological evaluation framework and does not involve clinician-led validation or aim to provide clinical decision support, as its objective is to assess explainability methods rather than clinical outcomes.

This work contributes to a growing body of benchmarking studies by explicitly aligning quantitative XAI evaluation with performance trade-offs commonly discussed in clinical tabular modeling contexts. Accordingly, the contribution lies at the framework level, formalization, controlled application, and reproducible comparison of established XAI methods, rather than in proposing new explainers or clinical decision rules.

The contributions of this work are:

- A *standardized, metric-driven evaluation framework* for explainable AI in healthcare tabular data, enabling controlled and reproducible comparison across heterogeneous XAI method families.
- Explicit mathematical operationalization and open-source implementation of commonly cited XAI evaluation metrics, enabling auditable and repeatable benchmarking rather than narrative comparison.
- A pre-specified focal-model selection protocol that controls model-induced variance in post-hoc XAI evaluation, isolating explainer effects.
- A unified empirical study spanning post-hoc attribution, rule-based explainers, and inherently interpretable models across multiple public healthcare tabular datasets.

#### Positioning relative to prior work

In contrast to prior surveys, toolkits, and evaluation studies [3–8], this work (i) formalizes widely used XAI metrics using explicit equations and open-source implementations, (ii) pre-specifies a single post-hoc explainee to control model-induced variability, (iii) evaluates post-hoc and ante-hoc methods within a single protocol, and (iv) benchmarks performance across multiple clinical tabular datasets. While prior work has highlighted the need for quantitative evaluation, existing studies remain fragmented: surveys largely provide conceptual definitions without unified operationalization, and toolkits such as Quantus focus on metric collections, primarily in neural network and image-based settings, without a controlled, end-to-end benchmarking protocol. In contrast, this work provides a unified instantiation of commonly cited metrics, explicitly tailored to clinical tabular data, integrating formalized equations, method–metric applicability constraints, and a pre-specified evaluation protocol within a single reproducible framework. A compact comparison is provided in Table 1.

**Table 1.** What this work adds vs. representative surveys/toolkits (✓= provided; – = not a primary focus).

| Survey/toolkit | Metric eqns | Open-source code | Pre-spec explaine | Post-hoc & ante-hoc | Multi-ds clin. tab. | Comment (vs. this work) |
| --- | --- | --- | --- | --- | --- | --- |
| Adadi & Berrada [3] | – | – | – | Post-hoc focus | – | Conceptual survey; no code/benchmarks. |
| Carvalho et al. [4] | Concepts | – | – | Both (survey) | – | Narrative notions; no formal equations/impl. |
| Zhou et al. [5] | Concepts | – | – | Both (survey) | – | Evaluation principles; no unified benchmark |
| Di Martino & Delmastro [6] | Survey | – | – | Mixed | – | Healthcare overview; not metric-focused |
| Hedström et al. [7] (Quantus) | Library | ✓ | – | Post-hoc | – | NN-centric metric library (image/saliency emphasis); no pre-specified explaine; no ante-hoc/rule-based; no clinical-tabular benchmark. |
| Nauta et al. [8] | Consistency notion | – | – | Post-hoc | – | Defines/analyzes consistency; not multi-metric benchmark |
| **This work** | ✓ | ✓ | ✓ | ✓ | ✓ | **Unified equations + open code; pre-specified explaine; post-hoc & ante-hoc; multi-dataset clinical tabular benchmark** |

Notes: “Metric eqns” = formal math definitions (not just verbal notions). “Pre-specified explaine” = rule-based protocol to fix a single post-hoc target model. “Multi-ds clin. tab.” = Multi-dataset clinical tabular

#### Scope

This paper presents a reusable *framework* for applying auditable, quantitative XAI metrics to clinical tabular tasks, including metric definitions/equations, implementation details, and method–metric applicability. Exhaustive coverage of all explainer families and parameter regimes is left for future extensions.

## Literature review

### The role of explainability in clinical AI

XAI is increasingly vital in healthcare, where decisions must be transparent, auditable, and trustworthy. As ML supports diagnosis, prognosis, and treatment planning, interpretability^1^ is essential for regulatory acceptance and practitioner trust [9, 10]. A wide spectrum of techniques exists, from instance-level feature attribution to global rule-based logic and visualization tools, aimed at rendering model behavior intelligible to clinicians [11–13]. XAI methods are often grouped into *post-hoc* and *ante-hoc* [14]. Post-hoc methods explain already-trained models; notable examples include SHAP (Shapley-value attributions), LIME (local surrogate explanations), and Anchors (precise local rules) [15–17]. Ante-hoc methods are inherently interpretable by design; examples include EBM (a GA^2^M with learned interactions) and TabNet (an attentive neural architecture with per-sample feature selection) [18, 19].

### XAI for tabular healthcare data

Despite their prevalence in clinical workflows (EHRs, registries, structured surveys), tabular datasets pose challenges such as missing values, class imbalance, and mixed data types [20]. Many healthcare XAI studies emphasize imaging and time series, or rely on proprietary data that limits reproducibility [1]. For tabular data specifically, comparative analyses include Hatwell et al. [21] (Ada-WHIPS, Anchors, LORE), Settouti et al. [22] (SHAP, LIME, permutation importance), and Alfeo et al. [23] (BoSCaR as a robust alternative to SHAP). Other works triangulate multiple explainers: Chadaga et al. [24] and Islam et al. [25] combine SHAP and LIME; Chun et al. [26], Khanna et al. [27], and Joseph et al. [28] cross-check attributions against known risk factors; Aiosa et al. [29] integrate SHAP with graphs; Peng et al. [30] pair SHAP with partial dependence. These examples illustrate growing use of XAI in clinical analytics, but most emphasize case-based interpretation rather than unified, metric-based comparisons across datasets.

#### Families

We distinguish (i) post-hoc feature-attribution methods, SHAP, which provides local Shapley values that can be aggregated to a global importance profile, and LIME, which is local by design; (ii) rule-based local explainers (Anchors); and (iii) inherently interpretable (ante-hoc) models (EBM, TabNet), both of which yield per-instance signals that can be aggregated to global views (e.g., EBM main-effect shapes; TabNet aggregated importances).

### Ensembles in clinical tabular machine learning

A recurring result in the tabular-ML literature is that tree-based ensembles are strong baselines on low- to mid-sized tabular datasets. Comparative studies report that gradient boosting and random forests frequently match or outperform linear models, kernel methods, and recent deep architectures on such data regimes [31, 32]. Two properties are repeatedly cited as drivers of this performance: (i) flexible handling of heterogeneous feature types and non-linear interactions, and (ii) robustness to routine preprocessing choices such as standard imputation. These characteristics are well-suited to clinical records (EHRs, registries, structured surveys) with varying feature scales and missing data.

Within explainability research, this prevalence has practical implications. Many clinical tabular pipelines adopt a high-performing tree ensemble and then attach a post-hoc local explainer (e.g., SHAP, LIME, Anchors) for per-instance interpretability, while inherently interpretable models (e.g., EBM and TabNet) are used when a transparent global structure is required. The literature thus motivates studying both families on tabular healthcare tasks rather than focusing on a single paradigm [1].

#### Takeaway

This literature motivates our emphasis on tree ensembles as competitive tabular baselines and, by extension, on post-hoc explainers commonly paired with them in practice.

### Quantitative XAI metrics

A recurring theme in the XAI literature is the call for *quantitative* criteria that move beyond narrative plausibility toward auditable evaluation [3, 4, 6, 33]. Several metrics have emerged as common currency, often defined conceptually in surveys and instantiated piecemeal in method papers or toolkits:

- **Fidelity.** Broadly, the agreement between an explanation and the behavior of the predictor being explained [4, 5]. Specific operationalizations include local accuracy for additive attributions (e.g., SHAP’s additivity guarantees and TreeSHAP’s exactness for tree ensembles) and surrogate–predictor agreement [15, 34].
- **Simplicity.** The cognitive compactness of the explanation, commonly proxied by sparsity (number of features/conditions) or length of the produced rationale [4, 35]. In rule learners, this often reduces to rule length; in attribution methods, thresholds on contribution magnitudes are frequently used.
- **Consistency (inter-explainer agreement).** Convergence across methods on what matters for the same instance/model, typically assessed via rank- or set-based concordance of salient features (e.g., correlations or overlap on top-*k* lists) [8].
- **Robustness.** Sensitivity of explanations to small, semantically irrelevant input changes; instantiated through input perturbations or sanity checks to ensure explanations reflect the model and data rather than artifacts [36, 37].
- **Precision and coverage for rule-based explanations.** For local if–then rationales (e.g., Anchors), empirical precision on covered instances and the proportion of instances covered are standard *rule-family evaluation metrics* [17].

While these metrics are widely recognized, their *operational forms* are scattered across individual papers, surveys, and general-purpose libraries. Survey articles largely provide conceptual definitions rather than unified, benchmark-ready formulations [3, 4, 6, 33], while recent studies have further highlighted the diversity of fidelity-oriented formulations and the lack of standardised evaluation protocols across settings [38]. Libraries such as Quantus collect implementations and tests, but are not tailored as end-to-end *clinical tabular* benchmarks with pre-specified explainee selection or a joint treatment of post-hoc and ante-hoc methods [7]. As a result, cross-paper comparisons often hinge on heterogeneous choices (datasets, predictors, thresholds, perturbation scales), making it difficult to assess trade-offs across methods. In line with recent surveys that frame properties such as faithfulness as high-level desiderata rather than single standardised metrics [39], we focus on concrete, measurable quantities that enable reproducible comparison across methods.

#### Canonical definitions in prior work

Several metrics already have operational mathematics in the literature: (i) additive local accuracy for SHAP/TreeSHAP with exact reconstruction on tree ensembles [34]; (ii) local surrogate agreement for LIME (weighted regression fit to *h* in a locality kernel) [16]; (iii) Anchors’ precision and coverage as rule-family metrics estimated against *h* [17]; (iv) rank-based agreement via Spearman/Kendall for top-*k* feature sets [8]; and (v) robustness via small input perturbations and explanation-distance sanity checks [36, 37].

#### Our instantiations vs. prior

Our contribution is not to introduce new metrics but to *standardize* one auditable instantiation per metric for clinical tabular settings and use them *side by side* in a single protocol: (1) *Fidelity* reported as both score- and decision-level; for SHAP/TreeSHAP we use exact additive reconstruction where applicable to avoid surrogate error. (2) *Simplicity* via a *relative* importance threshold (sparsity) so values are scale-free across methods/datasets. (3) *Consistency* as depth-averaged Spearman over top-*k* (we use *k*=5) to stabilize against ties and shallow lists. (4) *Robustness* as *ℓ*_1_ change under small Gaussian perturbations with fixed *σ* to make comparisons reproducible. (5) *Precision/Coverage* restricted to rule-family (Anchors) to avoid cross-paradigm conflation. Where multiple forms coexist in prior work, we justify our single choice for reproducibility and cross-dataset comparability. Formal equations and the method–metric applicability appear in *Materials & Methods*.

### Positioning relative to surveys and toolkits

Prior surveys synthesize concepts and taxonomies for explainability but stop short of a unified, auditable benchmarking recipe for clinical tabular settings [3, 4, 6, 33]. Method libraries such as Quantus assemble metric implementations and tests, yet they are not framed as end-to-end *benchmarks* with a pre-specified explainee selection protocol or a joint comparison of post-hoc and ante-hoc approaches on public clinical tabular data [7]. In contrast, we consolidate commonly cited metrics into explicit equations, release code, and apply them side-by-side across datasets within a single protocol that controls model-induced variance. A compact contrast with representative works appears in Table 1.

### Study design overview

We adopt a simple, auditable protocol tailored to clinical tabular data. First, we benchmark five predictors (GBC, RF, XGB, EBM, TabNet) on four datasets and pre-specify a single-model rule: select the focal explainee by best average F1 rank across datasets (accuracy as tie-breaker). Second, we compare two families of explainability within one setup: post-hoc explainers (LIME, SHAP/TreeSHAP, Anchors) are applied only to the selected predictor, while inherently interpretable models (EBM, TabNet) are analyzed via their native interpretability^2^. Third, we assess explanation quality with six established metrics, fidelity, simplicity, consistency, robustness, precision, and coverage, formalized with explicit equations and released as open-source code. Method-specific metric applicability is summarized in Table 2 (see *Materials and methods*); the overall protocol is illustrated in Fig. 1. We next detail datasets, models, and metric definitions in *Materials and methods*.

**Fig 1.**
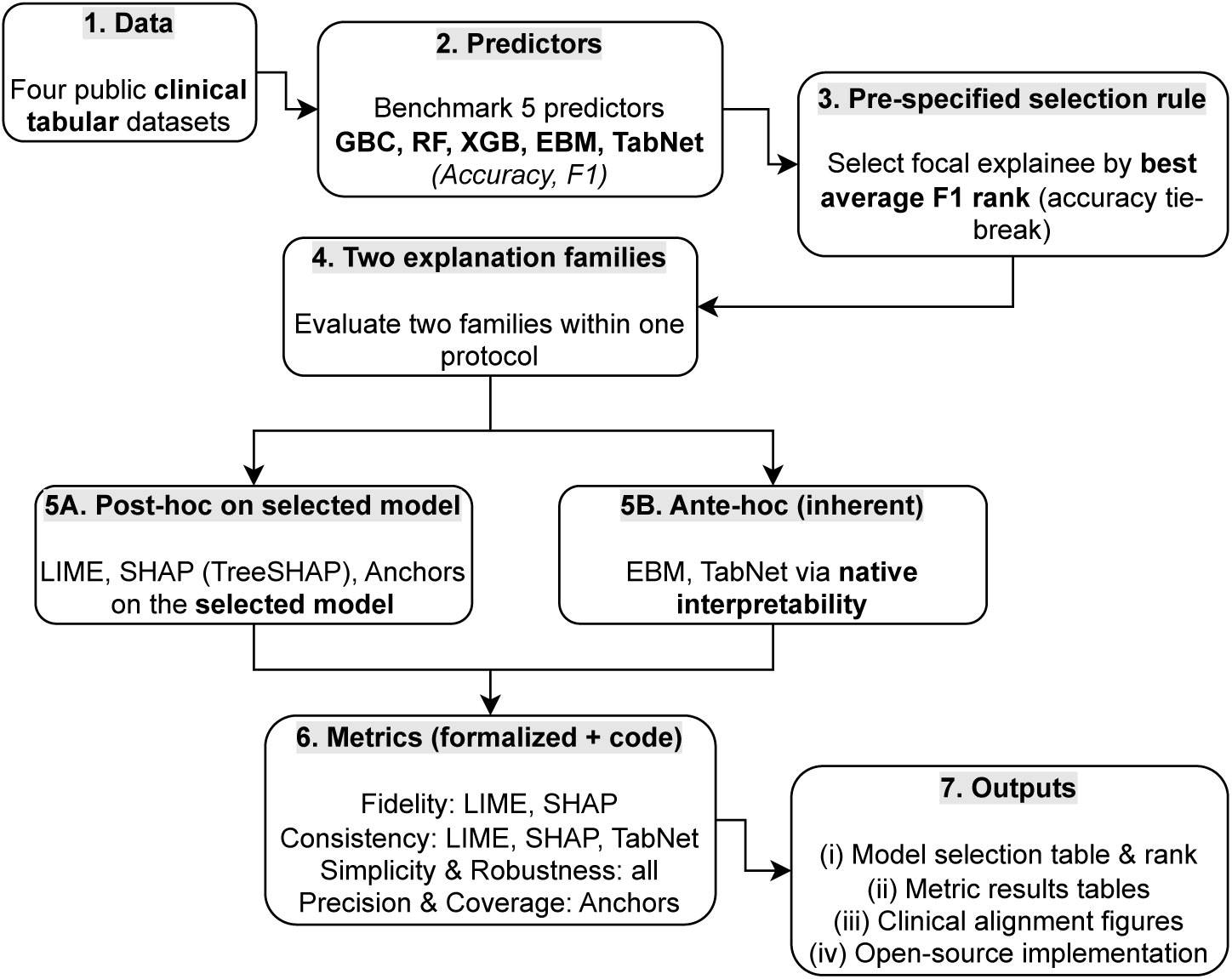
Study protocol at a glance. We benchmark five predictors on four clinical tabular datasets, pre-specify the focal explainee by best average F1 rank, apply post-hoc explainers (LIME, SHAP/TreeSHAP, Anchors) to that model, interpret ante-hoc models (EBM, TabNet) natively, and evaluate six metrics with method-specific applicability.

**Table 2.** Applicability of Evaluation Metrics to XAI Methods. All quantitative metrics are computed on *local* (per-instance) explanations; global summaries (e.g., aggregated SHAP importances, EBM main-effect plots, TabNet aggregated importances) are reported descriptively only. Metrics are applied only where theoretically meaningful: fidelity for surrogate-style local attributions (LIME; SHAP via TreeSHAP on trees), precision/coverage for rule learners (Anchors), rank-based consistency for methods producing single-feature rankings (LIME, SHAP, TabNet), and simplicity/robustness across families.

| Evaluation Metric | LIME | SHAP | Anchors | EBM | TabNet |
| --- | --- | --- | --- | --- | --- |
| **Fidelity** | ✓ | ✓ | - | - | - |
| **Simplicity** | ✓ | ✓ | ✓ | ✓ | ✓ |
| **Consistency** | ✓ | ✓ | - | - | ✓ |
| **Robustness** | ✓ | ✓ | ✓ | ✓ | ✓ |
| **Precision** | - | - | ✓ | - | - |
| **Coverage** | - | - | ✓ | - | - |

## Materials and methods

### Ethics statement

This study used publicly available, anonymized datasets and did not involve human subjects or personally identifiable data. No new data was collected, and all analyses prioritized research integrity and reproducibility. All datasets were accessed under their respective licenses/terms of use.

### Scope and rationale for method selection

We compare two complementary families of explainability on clinical tabular tasks: *local post-hoc* explainers applied to a fitted predictor (LIME, SHAP/TreeSHAP, Anchors) and *ante-hoc* models whose structure is itself interpretable (EBM, TabNet). These choices are guided by prior work [1, 4, 5] and by:

a) widespread adoption in healthcare ML pipelines for tabular data,
b) conceptual diversity (attribution, local rules, and glass-box models), and
c) coverage of established quantitative metrics (simplicity and robustness across families; fidelity for surrogate-style local attributions; consistency where ranked single-feature importances exist; precision and coverage for rule-based Anchors).

Concretely, we selected one representative per major explainer family commonly used in tabular healthcare pipelines (feature attribution, local rules) and paired them with two widely used interpretable-by-design baselines (EBM and TabNet) to cover both post-hoc and ante-hoc paradigms under a single protocol.

To avoid confounding explainer comparisons with model changes, all post-hoc methods are applied to a single, pre-specified focal predictor chosen by the best average F1 rank across datasets (accuracy as tie-breaker). Ante-hoc models are assessed via their native interpretability rather than as post-hoc targets. This design supports controlled comparison within the post-hoc subset, while Table 2 makes the broader method–metric applicability explicit.

All quantitative evaluation metrics in this study are applied to *local* (per-instance) explanations; global aggregates (e.g., mean |SHAP|, EBM shape functions) are reported for context and are not included in metric scoring.

#### Global RF baselines

For the focal Random Forest, we also compute two standard global feature importances, permutation importance (PFI) and mean decrease in impurity (Gini/MDI). These provide conventional tree-model baselines to triangulate global rankings against SHAP/EBM/TabNet summaries (shown in the Appendix) and are not part of the local-metric scoring.

##### Alternatives and scope

The rule-based space includes other candidates (e.g., rule lists/sets). Interpretable modeling also spans linear/monotone GAM variants and decision sets. We do not claim exhaustiveness; rather, we provide a reproducible, metric-driven baseline with widely used representatives and release code to ease extension. Counterfactuals and example-based methods are orthogonal families we list for future work (see *Limitations*).

### Datasets

Four publicly available healthcare tabular datasets were selected using the following inclusion criteria: each was publicly accessible, non-synthetic, suitable for binary classification, and either contained no missing values or had missing values already imputed in the publicly released version (i.e., no additional imputation by us).

1. **PDHS (C-section)**: Contains 26,284 rows and 24 columns. Derived from national health surveys; the publicly released version includes imputed values for originally missing entries. Used to predict Caesarean-section outcomes [25].
2. **Diabetes retinopathy**: Includes 1,151 rows and 19 columns. Used to assess the risk of diabetes retinopathy progression [21].
3. **Breast cancer**: Comprises 569 rows and 30 columns. Applied for binary classification of breast cancer diagnoses [21].
4. **ESDRPD (Early Stage Diabetes Prediction)**: Contains 520 rows and 16 columns. Targets the early detection of diabetes-related complications [28].

All tasks are binary classification using the dataset’s default target variable.

### Machine learning models

We evaluated five machine learning models: Gradient Boosting Classifier (GBC), Random Forest (RF), Extreme Gradient Boosting (XGB), TabNet, and Explainable Boosting Machines (EBM). Model performance was assessed using accuracy and F1 score to establish a predictive baseline across different methodological families.

Our emphasis on tree-based ensembles (RF, GBC, XGB) reflects both the structure of the datasets and converging evidence from the literature. All four datasets used in this study are low- to mid-sized tabular healthcare datasets (ranging from 520 to 26,284 rows, with mixed categorical and numerical features). For such data regimes, ensemble-based tree methods have repeatedly been shown to outperform linear models, kernel methods, and even recent deep learning architectures for tabular data [31, 32]. They handle heterogeneous predictors, capture non-linear interactions, and work well with standard imputation, properties that are particularly relevant to clinical data. The recent survey of explainable AI in healthcare similarly found that tree ensembles are among the most frequently reported “winner” models in comparative studies [1]. Evidence from other domains aligns with this conclusion, e.g., systematic churn-prediction reviews where ensembles dominate in tabular data settings [40].

To complement these ensembles, we also included two inherently interpretable models: EBM and TabNet. EBM is a generalized additive model with automatic interaction detection, inspired by boosting methods but designed as a glass-box approach in which feature effects and interactions can be inspected directly without post-hoc explanations [41]. TabNet is a deep learning architecture specifically developed for tabular datasets; it employs sequential attention masks to select salient features for each sample, thereby providing feature-level interpretability alongside competitive predictive performance [19]. Including these models ensured that both inherently interpretable and neural architectures were represented in the evaluation, not only ensemble-based methods.

### XAI methods and evaluation metrics

Concrete implementation details, software packages, and environment specifications are summarized in the *Experimental setup* to support exact reproducibility.

#### Methods policy

This policy reflects the principle, supported in prior XAI literature, that evaluation metrics must be aligned with the structural form and semantics of the explanation, for example numeric attribution vectors versus logical rules, rather than applied uniformly across heterogeneous paradigms [4, 8, 17]. We compute all metrics on *local* (per-instance) outputs with fixed parameters and seeds; apply each metric only where it is theoretically meaningful for the explainer family; and run all post-hoc explainers on a single, pre-specified focal predictor to ensure controlled comparability across datasets.

We evaluated five XAI methods spanning post-hoc and inherently interpretable approaches: LIME, SHAP^3^, Anchors, EBM, and TabNet. These methods are selected to represent three complementary paradigms, post-hoc attribution, rule-based local explanations, and inherently interpretable models, providing conceptual coverage rather than method-specific focus. This selection is therefore principled and non-arbitrary, ensuring representation of distinct XAI paradigms within a single evaluation protocol. To isolate differences attributable to explainers, all post-hoc methods (LIME, SHAP, Anchors) are applied to a single focal predictor chosen after benchmarking in the *Results* section. In contrast, EBM and TabNet are assessed via their inherent (ante-hoc) interpretability.

We use six established quantitative evaluation metrics from prior work: *fidelity*, *simplicity*, *consistency*, *robustness*, *precision*, and *coverage*. Although the metrics are not new, we formalize and implement their mathematical definitions for transparency and reproducibility [4, 5, 8]. Fidelity applies to post-hoc feature-attribution surrogates (LIME, SHAP); precision and coverage apply to rule-based explainers (Anchors); simplicity and robustness apply to all methods; and consistency is reported for methods that yield ranked single-feature importances^4^ (LIME, SHAP, TabNet). Table 2 defines an explicit method–metric applicability mapping, ensuring that each metric is used only where theoretically valid given the structure of the explanation, and preventing invalid cross-paradigm comparisons.

#### Parameter choices and sensitivity

Several evaluation metrics require operational parameters, including the simplicity threshold *τ*, the top-*k* depth used for rank-based consistency, and the perturbation scale *σ* for robustness analysis. Rather than tuning these parameters to optimize results on a specific dataset, we adopt representative and conservative settings that are widely used in empirical XAI evaluation to characterize explanation behavior under standard operating conditions.

Specifically, simplicity thresholds *τ* ∈ {0.10, 0.05, 0.01} are used to examine explanation sparsity across coarse-to-fine attribution cutoffs. This follows prior work that evaluates simplicity using relative importance thresholds and reports sparsity trends across multiple operating points, reflecting the absence of a canonical sparsity cutoff and the context-dependent nature of explanation compactness [35, 44].

For rank-based consistency, we fix *k* = 5 and compute agreement over the top-ranked features only, focusing on the most salient elements of each explanation. This choice aligns with comparative XAI studies that restrict consistency analysis to a small number of dominant features to balance interpretability and stability, and to avoid noise introduced by low-importance feature tails [5, 8, 15, 16].

For robustness, we use a small perturbation scale *σ* = 0.01, introducing element-wise Gaussian noise as a fixed numerical sensitivity probe. The purpose is to assess how explanation outputs change under a reproducible perturbation protocol that can be applied consistently across datasets and explainer families. Because the perturbation is applied to the numerically encoded input representation, including integer-coded fields, it does not enforce feature-specific codebooks, categorical-level constraints, ordinal structure, or physiological ranges. Accordingly, robustness scores are interpreted as explanation sensitivity under Gaussian jitter, rather than as evidence of stability under clinically meaningful patient variation [7, 37, 45].

Partial sensitivity analysis is reflected in the reported results: simplicity is evaluated across multiple *τ* values, and Anchors are assessed across multiple precision thresholds. A more exhaustive sensitivity study over {*k, σ*} and alternative perturbation models would be valuable but is beyond the scope of the present work and is identified as future research (see *Limitations and Future Work*).

#### Metric definitions and relation to prior work

Our fidelity follows the standard *agreement-based* notion in the XAI literature, i.e., how closely an explainer or surrogate reproduces the output of the predictive model for the same input [4, 5]: for LIME we compare surrogate outputs to the predictor; for SHAP we reconstruct the predictor’s output from the additive explanation (local accuracy), which is exact for tree ensembles under TreeSHAP [34]. Our simplicity counts features above a relative-importance threshold [35]. Consistency uses Spearman correlation over top-*k* ranked features (agreement across explainers) [8]. Robustness measures the sensitivity of local explanations to small input perturbations [36]. For Anchors, we report rule precision and coverage as in the original work [17]. Our contribution is to operationalize these notions with unified equations and open-source code for comparative, multi-dataset evaluation in clinical tabular settings.

We now present metric definitions for evaluating the two classes of explainers: feature attribution–based (e.g., SHAP, LIME) and rule-based (e.g., Anchors). All metrics are defined over a dataset *X* = {*x*_1_*, x*_2_*, …, x_N_* } of *N* instances, unless stated otherwise.

#### Common notation

Let 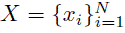 be the dataset, *F* the set of features (with |*F* | elements), *h* the trained predictor, and *M_h_* an explainer instantiated with respect to *h*. We write *P_h_*(*x*) for the predictor’s score and 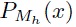 for the explainer’s surrogate/reconstruction score when defined (e.g., LIME, TreeSHAP). For local explanations on *x_i_* we use *E*(*M_h_, x_i_*) to denote the explanation object: (i) for attribution methods, 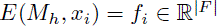 with entries *f_ij_*; (ii) for rule-based methods, *E*(*M_h_, x_i_*) = *A_i_* ⊆ *F* is the set of feature conditions (literals) in the rule.

### Evaluating feature importance-based XAI methods

We formalize established quantitative metrics for attribution-based explainers below.

#### Fidelity

Fidelity [46] evaluates how accurately an XAI method replicates the behavior of the original predictive model. It is computed in two complementary ways:

- **Score fidelity:** Measures the similarity between the model’s probability and the explainer’s reconstruction, operationalized as 1 − *P_h_*(*x_i_*) − *P_Mh_* (*x_i_*).
- **Decision fidelity:** Measures the agreement in final class predictions between the predictive model and an explainer-induced approximation (e.g., a local surrogate model where applicable). It reflects how often the label derived from the explainer’s approximation matches the label of the original model. This follows standard definitions of fidelity as agreement between a model and its explanation approximation [38].

The formal equations are:

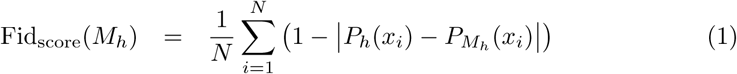

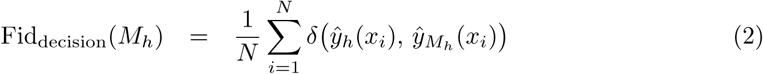

Where:

- *P_h_*(*x_i_*) is the probability (score) from the trained predictor *h* at *x_i_*.
- *P_Mh_* (*x_i_*) is the surrogate/reconstruction probability produced by the explainer *M_h_*.
- 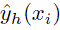 and 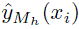 are the corresponding predicted class labels from *h* and *M_h_*.
- *δ*(*a, b*) is the Kronecker delta, returning 1 if *a* = *b* and 0 otherwise.

This dual view of fidelity allows both probabilistic closeness and class decision alignment to be assessed, highlighting whether an explanation preserves not just the model’s outcome but its internal confidence as well.

A perfect score of 1.0 indicates that the explainer exactly replicates the model’s predicted probability (score fidelity) or predicted label (decision fidelity); small deviations can arise from ties or probabilistic rounding. Scores closer to 0.0 signify poor alignment between the explainer and the model.

#### Simplicity (attribution-based)

Simplicity [35] quantifies how concise and interpretable an explanation is, based on the number of features it includes. In high-stakes domains like healthcare, simpler explanations involving fewer features are often preferred to support user trust and rapid decision-making.

For attribution methods, we define simplicity as the *average number of features per instance* whose relative importance exceeds a threshold *τ* ∈ {0.10, 0.05, 0.01}. Let *f_i_* = [*f_i_*_1_*, …, f_i_*_|*F* |_] be the attribution vector for instance *x_i_*. Features with 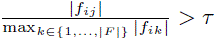 are counted toward the explanation.

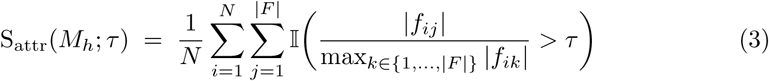

Where:

- *F*: set of features (with cardinality |*F* |).
- *f_ij_*: attribution score of feature *j* for instance *x_i_*.
- max*_k_*_∈{1*,…,*|*F* |}_ |*f_ik_*|: maximum absolute attribution in instance *x_i_*.
- *τ* is the threshold value ∈ {0.10, 0.05, 0.01}.
- Ι(·): indicator function, returning 1 if the condition is true, 0 otherwise.

The threshold parameter controls explanation sparsity. Higher thresholds (e.g., 0.1) retain only the most influential features, resulting in simpler, more focused explanations. Lower thresholds (e.g., 0.01) capture weaker attributions, producing denser explanations with more features.

This formulation enables consistent comparison of explanation conciseness across methods and datasets. Lower values indicate simpler explanations involving fewer features, which are generally easier for human interpretation. Higher values may capture more detail but reduce interpretability.

#### Consistency

Consistency [8] measures the extent to which explanation methods assign similar relative importance to the same features for a given instance. We use Spearman’s rank correlation because the objective is to compare feature-order agreement rather than absolute attribution magnitudes.

Pairwise consistency is computed across progressive feature depths. For each instance, the top *n* ranked features from two methods are compared for *n* = 2*, …, k*, where *k* = 5. The case *n* = 1 is excluded because rank correlation is not defined for a single ranked item. This formulation focuses on agreement across the most salient part of the explanation while avoiding instability from low-importance feature tails.

The consistency score between two XAI methods *M*_1*,h*_ and *M*_2*,h*_ is defined as:

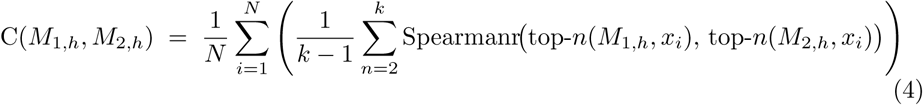

Where:

- top-*n*(*M*_1*,h*_*, x_i_*) and top-*n*(*M*_2*,h*_*, x_i_*): denote the top-*n* ranked feature lists for instance *x_i_* from methods *M*_1*,h*_ and *M*_2*,h*_, with feature identities aligned prior to computing the correlation.
- Spearmanr(·, ·): denotes Spearman’s rank correlation.
- *k* = 5: the maximum number of top-ranked features considered.

The resulting score lies in the range [−1, 1], where larger positive values indicate stronger agreement in feature ranking, values near zero indicate weak or no rank agreement, and negative values indicate opposing ranking patterns. For comparisons involving TabNet, the score should be interpreted as cross-configuration rank agreement because TabNet explanations are obtained from the native TabNet model, whereas LIME and SHAP explain the selected RF focal predictor.

#### Robustness (attribution-based)

Robustness [36] evaluates how stable a feature-based explanation remains when small input perturbations are introduced. In this study, it is operationalized using the fixed Gaussian-jitter protocol described above.

We ntroduce Gaussian noise to each instance, drawn from N (0*, σ*^2^) with *σ* = 0.01, applied element-wise to the numerically encoded input representation with no model refitting. Robustness R_attr_(*M_h_*) is computed as the average *ℓ*_1_ change between explanation vectors before and after a small perturbation^5^:

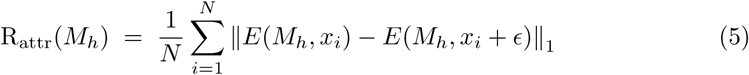

Where:

- *ɛ* ∼ N (0*, σ*^2^): element-wise perturbation.
- *E*(*M_h_, x_i_*): explanation vector from method *M_h_* for input *x_i_*.

Lower values indicate higher robustness, i.e., explanations remain stable even under small input perturbations. A value of 0 means the explanation is entirely unchanged. These values therefore quantify explanation sensitivity under the benchmark perturbation protocol.

#### Transition to rule-based explainers

The metrics discussed thus far apply to explainers that yield *numeric, per-instance feature-importance vectors*. In contrast, rule-based explainers such as Anchors produce logical decision rules, for which precision and coverage are native evaluation criteria [17]. This necessitates a distinct set of metrics tailored to symbolic conditions and coverage-based evaluation. We now describe four such metrics used to evaluate rule-based explanations.

### Evaluating rule-based XAI methods

#### Notation reminder (rule-based)

In this subsection *E*(*M_h_, x_i_*) = *A_i_* ⊆ *F* denotes the set of feature conditions (literals) in the rule for *x_i_*; primes (e.g., *A*^′^) indicate the rule after a small perturbation of *x_i_*.

Rule-based XAI methods, such as Anchors, generate explanations in the form of logical if-then rules composed of discrete feature conditions (e.g., “*Age >* 40 AND *Gender* = *Female*”). These rules differ fundamentally from numeric feature attributions and thus require tailored evaluation metrics. We define four such metrics: **precision**, **coverage**, **simplicity**, and **robustness**, detailed below.

##### Precision and coverage

*Precision* measures how often the black-box predictor agrees with the explained-instance prediction on the samples covered by the rule, and *coverage* is the fraction of instances to which the rule applies [17]. Tightening the precision threshold typically increases reliability while reducing coverage (the canonical precision–coverage trade-off).

Let 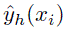 be the prediction of *h* on the explained instance *x_i_*, and let Ι[·] be the indicator. Then

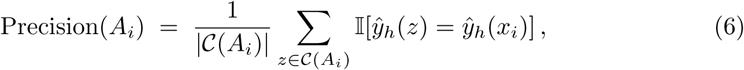

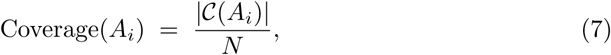

Where:

- *z* denotes a generic instance in the dataset *X*.
- C(*A_i_*) = {*z* ∈ *X*: *A_i_* matches *z*}.
- *N* is the dataset size used for coverage.

These metrics are computed using the Anchor package’s inbuilt estimators and reflect how reliably and broadly each generated rule captures the model’s behavior.

A perfect precision score is 1.0 (i.e., 100% correct on all covered instances). Coverage values range from 0 to 1, where 1 indicates full dataset applicability.

Higher thresholds (e.g., 0.99) enforce stricter rule precision, producing explanations with near-perfect reliability. However, this typically reduces coverage, as the rules apply to fewer data instances. This trade-off is particularly relevant in healthcare: while high-precision rules improve trust in local predictions, their narrow applicability must be weighed against the need for generalizability across diverse patient groups.

##### Simplicity (rule length)

Simplicity equals rule length (number of literals)^6^.

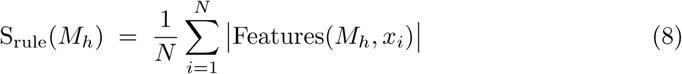

Where:

- Features(*M_h_, x_i_*): the set of features used in the rule that explains *x_i_*.

A smaller simplicity score may be desirable in clinical settings to support quick comprehension and decision-making.

##### Robustness (rule-based)

Robustness for rule-based explanations reflects the stability of the rule features under the same fixed perturbation protocol. If different rules are generated from perturbed inputs, this indicates lower rule stability. We introduce small Gaussian noise to the input instance, drawn from N (0*, σ*^2^), with *σ* = 0.01, before recomputing the rules.

Following the same perturbation approach as attribution methods [36], we measure robustness via the Jaccard dissimilarity^7^ between pre- and post-noise rule feature sets, as defined in Eq. (9).

Let *A_i_* be the set of features used in the rule for instance *x_i_*, and *A*^′^ the rule after applying small Gaussian noise to the input *x_i_*. Then:

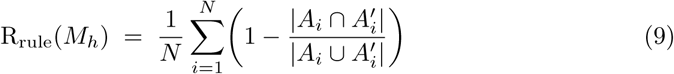

Where:

- *A_i_* is the set of feature conditions in the rule for *x_i_*, and *A*^′^ is the corresponding set after a small perturbation is applied to *x_i_* (i.e, *x_i_* + *ɛ*).

Lower scores indicate greater stability of the rule structure under the fixed perturbation protocol. A robustness score of 0.0 means the rule remains identical after perturbation.

#### Framework components and extensibility

The framework comprises: (i) explicit equations for six established metrics; (ii) a pre-specified focal-model protocol to control model-induced variance; (iii) a metric–method applicability map; and (iv) open-source code. New explainers or metrics can be added by implementing the metric interface (inputs/outputs documented in the repository) and extending the applicability map; the protocol remains unchanged.

### Experimental setup

Each dataset was partitioned into training and testing subsets by preserving original splits where available or applying a standard 70:30 split otherwise. The training sets were used to fit all five models (GBC, RF, XGB, TabNet, EBM). Predictive performance was evaluated using accuracy and F1 score on the test sets.

To control for model-induced variability in the post-hoc explainability study, we pre-specified a single-model protocol: after benchmarking, we select one tree-ensemble as the focal explainee using an aggregate performance rule. The rule chooses the model with the best *average rank across datasets* on F1 (primary) and accuracy (tie-breaker); if models are tied, we prefer the one with native support for exact local attributions to minimize explainer-approximation error. Post-hoc explainers (LIME, SHAP, Anchors) are then computed on the selected focal model, while EBM and TabNet are examined via their inherent interpretability.

#### Why a single explainee?

Fixing one competitive predictor for post-hoc comparisons controls model-induced variance so that differences are attributable to the explainer rather than to changes in the underlying decision function [7, 37]. Selecting a different best-performing predictor for each dataset, or applying all post-hoc explainers to all candidate predictors, would introduce an additional experimental axis and shift the study from controlled metric evaluation toward explainer–model interaction analysis. This latter question is important but outside the scope of the present framework demonstration.

##### Hyperparameter policy and reproducibility

Because our primary goal is to benchmark *explainability*, not to maximize predictive accuracy, we adopt library defaults with a fixed global seed and avoid extensive per-dataset hyperparameter optimization. This standardization reduces model-induced variability so that observed differences are attributable to the explainers themselves, in line with recent guidance on rigorous and transparent XAI evaluation and reproducible ML workflows [7, 47, 48].

Moreover, explanation quality is known to be sensitive to modeling and procedural choices, which motivates controlling additional sources of variance introduced by heavy hyperparameter optimization [37]. For Random Forests specifically, prior work shows diminishing returns once the number of trees is sufficiently large and argues that treating the tree count as a major tuning parameter is often unnecessary; performance typically plateaus around ∼100–200 trees [49, 50]. Accordingly, we use defaults with a fixed seed for all models and a conventional tree count for RF.^8^

##### Software environment and packages

All experiments were conducted in Python 3.11.7. Explainability methods were implemented using established open-source libraries. Specifically, LIME explanations were generated using the lime, SHAP values were computed using the shap, rule-based explanations were generated using the anchor-exp, EBM were implemented via interpret, and TabNet models were trained using the pytorch-tabnet package. The full software environment, including all Python dependencies, was captured using pip freeze and is released as a requirements.txt file alongside the source code to support exact reproducibility.

## Results

### Classification performance

We benchmarked five classifiers (GBC, RF, XGB, TabNet, EBM) across the four healthcare datasets. Table 3 reports the accuracy and F1 scores for each model–dataset pair.

**Table 3.**
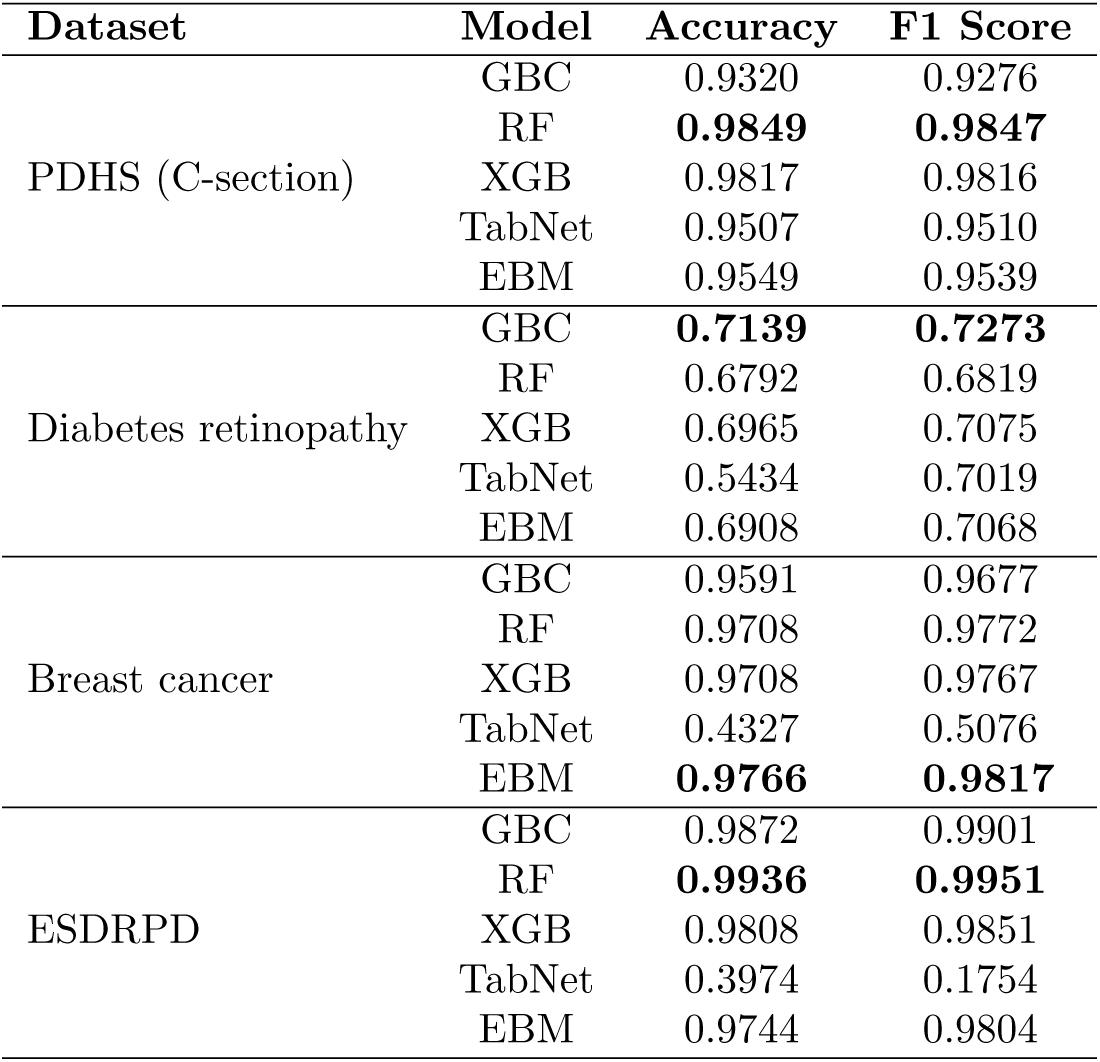
Classification performance (Accuracy, F1) across datasets; best per dataset in **bold** (primary: F1; tie-breaker: Accuracy).

#### Role of this benchmark

Table 3 is the input to our pre-specified focal-model protocol: we report per-dataset performance so that ranks and the overall *average F1 rank* can be computed transparently.

#### Model selection rule (pre-specified)

As outlined in the *Experimental setup*, the single focal explainee for post-hoc XAI is selected by *best average rank on F1 across datasets* (primary), with *average rank on accuracy* as a tie-breaker if needed.

#### Selection outcome

Applying the rule yields “Random Forest (RF)” as the focal post-hoc explainee (see Table 4): RF ranks first on two datasets (PDHS, ESDRPD), second on Breast cancer, and fifth on Diabetes retinopathy, giving the best average F1 rank (2.25). The accuracy tie-breaker was not required. RF is not the best-performing classifier on every dataset, particularly Diabetes retinopathy. The focal-model protocol therefore selects a competitive aggregate explainee for controlled post-hoc comparison, rather than a per-dataset optimal classifier.

**Table 4.** Per-dataset F1 ranks (1 = best) and average rank used for model selection.

| Model | PDHS<br>(C-section) | Diabetes<br>retinopathy | Breast<br>cancer | ESDRPD | Avg. rank<br>(F1) |
| --- | --- | --- | --- | --- | --- |
| RF | **1** | 5 | 2 | **1** | **2.25** |
| XGB | 2 | 2 | 3 | 3 | 2.50 |
| EBM | 3 | 3 | **1** | 4 | 2.75 |
| GBC | 5 | **1** | 4 | 2 | 3.00 |
| TabNet | 4 | 4 | 5 | 5 | 4.50 |

#### Implication for XAI

Having selected RF by a pre-specified, data-driven rule, we compute post-hoc explanations (SHAP, LIME, Anchors) on RF to control for model-induced variability. EBM and TabNet are reported via their inherent interpretability.

### Evaluation of XAI methods aggregated over local explanations

The explanation outputs were assessed using predefined metrics: fidelity, simplicity, consistency, robustness, precision, and coverage. The results, averaged over local instance explanations, summarize explanation behavior across the different datasets. The XAI results should be interpreted according to the method–metric applicability design in Table 2. LIME, SHAP, and Anchors explain the selected RF focal predictor, whereas EBM and TabNet are evaluated through their native model-specific interpretability mechanisms. Thus, mixed-family comparisons involving EBM or TabNet describe method–predictor configurations rather than explanations of a common underlying predictive function. These results should therefore be read alongside the corresponding predictive performance in Table 3.

#### Evaluation of feature importance-based XAI methods

##### Fidelity

The fidelity scores for LIME and SHAP are reported in Table 5. In this context, fidelity is assessed using two complementary criteria: *Score* fidelity, which measures agreement in predicted probabilities, and *Decision* fidelity, which measures agreement in predicted class labels.

**Table 5.**
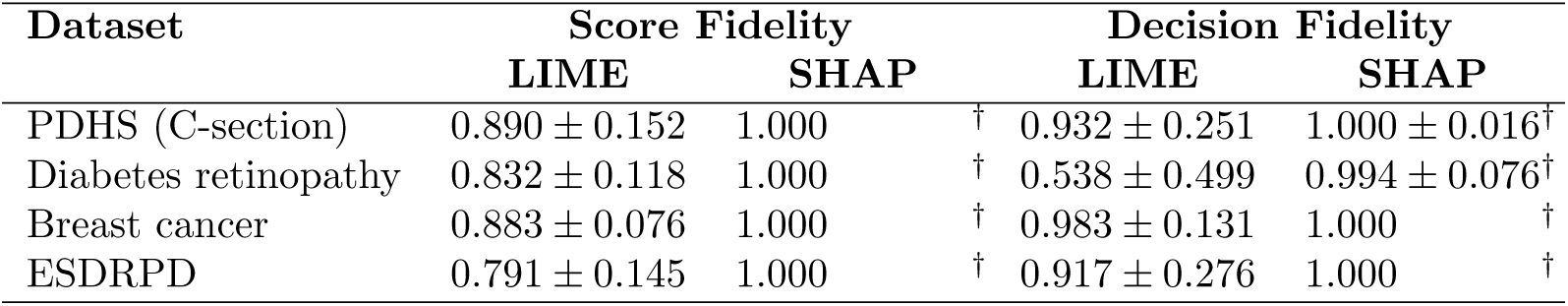
Fidelity of LIME and SHAP explanations, reported as mean ± standard deviation over test instances. The highest numerical values are marked with ^†^; for SHAP (TreeSHAP on tree ensembles), score fidelity of 1.000 reflects exact additive reconstruction rather than a comparative surrogate approximation outcome. Standard deviations are not shown when they round to 0.000.

SHAP, using TreeSHAP, obtained score fidelity of 1.000 across all datasets with zero variance. This reflects the local accuracy property of TreeSHAP for tree ensembles, where additive feature attributions exactly reconstruct the model output [15, 34]. It should therefore be interpreted as an expected property of the RF–TreeSHAP pairing, rather than as an empirical superiority result under comparable surrogate-approximation conditions. In contrast, LIME’s fidelity reflects the quality of a local surrogate approximation and can vary across instances. Minor deviations in decision fidelity can still occur due to thresholding and rounding effects when converting probabilities into class labels (e.g., 0.994 ± 0.076 for Diabetes retinopathy), despite near-identical probability reconstruction.

LIME generally exhibited lower fidelity and substantially higher variability across instances. For example, in the Diabetes retinopathy dataset, LIME achieved a score fidelity of 0.832 ± 0.118 and a markedly lower decision fidelity of 0.538 ± 0.499, indicating unstable local approximations at the instance level. Comparable patterns are observed in ESDRPD (0.791 ± 0.145 for score fidelity and 0.917 ± 0.276 for decision fidelity), highlighting LIME’s sensitivity to complex or non-linear decision boundaries. These findings underscore that LIME’s linear approximation of decision boundaries often sacrifices an accurate reflection of model behavior.

##### Simplicity

The simplicity of LIME, SHAP, EBM, and TabNet is measured by the number of features involved in the explanations, as reported in Table 6. Results are summarized using mean ± standard deviation over test instances to capture both the typical explanation size and its variability across instances.

**Table 6.** Simplicity of XAI method–predictor configurations, measured as the number of features in the explanation and reported as mean ± standard deviation over test instances. LIME and SHAP explain the selected RF focal predictor, whereas EBM and TabNet are evaluated through native model-specific interpretability. Lowest values are marked descriptively with ^†^ and should be interpreted alongside the corresponding predictive performance in Table 3. Standard deviations are not shown when they round to 0.000.

| Method | PDHS<br>(C-section) | Diabetes<br>retinopathy | Breast<br>cancer | ESDRPD |
| --- | --- | --- | --- | --- |
| **Threshold = 0.1** |  |  |  |  |
| LIME | $1.82 \pm 0.39^\dagger$ | $9.86 \pm 0.42$ | $9.93 \pm 0.37$ | $9.95 \pm 0.22$ |
| SHAP | $3.86 \pm 2.82$ | $11.44 \pm 2.52$ | $13.86 \pm 2.44$ | $8.93 \pm 1.99^\dagger$ |
| EBM | $7.68 \pm 3.93$ | $20.11 \pm 5.95$ | $28.40 \pm 5.37$ | $20.53 \pm 2.12$ |
| TabNet | $4.31 \pm 1.38$ | $6.10 \pm 7.07^\dagger$ | $6.21 \pm 9.56^\dagger$ | $11.53 \pm 6.46$ |
| **Threshold = 0.05** |  |  |  |  |
| LIME | $3.27 \pm 0.79^\dagger$ | $10.00 \pm 0.05$ | 10.000 | 10.000 $^\dagger$ |
| SHAP | $8.59 \pm 3.37$ | $14.23 \pm 2.06$ | $17.42 \pm 2.83$ | $12.28 \pm 1.46$ |
| EBM | $16.26 \pm 7.08$ | $27.16 \pm 4.55$ | $43.55 \pm 5.20$ | $27.74 \pm 1.92$ |
| TabNet | $4.85 \pm 1.39$ | $7.34 \pm 7.53^\dagger$ | $6.69 \pm 9.73^\dagger$ | $12.33 \pm 5.74$ |
| **Threshold = 0.01** |  |  |  |  |
| LIME | 10.000 | 10.000 | 10.000 | 10.000 $^\dagger$ |
| SHAP | $18.09 \pm 1.93$ | $17.10 \pm 1.14$ | $25.46 \pm 2.34$ | $15.37 \pm 0.75$ |
| EBM | $35.96 \pm 4.83$ | $35.63 \pm 1.79$ | $56.08 \pm 1.56$ | $30.62 \pm 0.98$ |
| TabNet | $5.46 \pm 1.46^\dagger$ | $9.88 \pm 7.64^\dagger$ | $7.37 \pm 9.84^\dagger$ | $13.49 \pm 4.13$ |

The thresholds (0.1, 0.05, and 0.01) represent different levels of feature-importance cutoffs used to determine which features are included in an explanation. Specifically, the threshold indicates a fraction of the maximum absolute feature-attribution value in a given instance. A feature is included in the explanation if its absolute value exceeds the threshold multiplied by the maximum absolute value in that instance. Higher thresholds therefore result in fewer features and simpler explanations, while lower thresholds admit more features and yield more detailed explanations. Because the maximum absolute feature value varies across datasets and instances^9^, the resulting explanation sizes also vary, as formalized in Eq. (3).

Within the method–predictor interpretation described above, TabNet frequently produced compact native explanations, particularly at lower thresholds (0.05 and 0.01). However, these compactness results should not be interpreted independently of predictive performance. In particular, TabNet’s simplicity values on Breast cancer and ESDRPD should be read cautiously given its weak classification performance on those datasets in Table 3. LIME produced compact RF-based local explanations in some settings and, at lower thresholds, often converged to a fixed number of selected features with near-zero deviation. SHAP explanations were generally denser, while EBM generated more feature-rich native explanations across thresholds.

The reported standard deviations highlight important differences in instance-level behavior. TabNet and SHAP exhibit higher variability in explanation size, reflecting adaptive feature selection that can change substantially across instances, whereas LIME often produces explanations of consistent size once the threshold constraint becomes binding. Overall, these results describe an explanation compactness–detail trade-off across the evaluated method–predictor configurations, rather than a model-independent ranking of XAI methods.

##### Consistency

The consistency scores between LIME, SHAP, and TabNet are reported in Table 7. Results are summarized using mean ± standard deviation over test instances to capture both average agreement and instance-level variability in feature rankings.

**Table 7.** Consistency between explanation-producing configurations measured using Spearman rank correlation over *n* = 2*, …, k*, with *k* = 5, reported as mean ± standard deviation over test instances. LIME and SHAP explain the selected RF focal predictor, whereas TabNet rankings are obtained from the native TabNet model. Highest values within each dataset are marked descriptively with ^†^.

| Dataset | LIME–SHAP | LIME–TabNet | TabNet–SHAP |
| --- | --- | --- | --- |
| PDHS (C-section) | $0.46 \pm 0.23^\dagger$ | $-0.08 \pm 0.25$ | $-0.06 \pm 0.27$ |
| Diabetes retinopathy | $0.08 \pm 0.49^\dagger$ | $-0.50 \pm 0.28$ | $-0.47 \pm 0.32$ |
| Breast cancer | $0.16 \pm 0.42^\dagger$ | $-0.52 \pm 0.33$ | $-0.50 \pm 0.29$ |
| ESDRPD | $0.66 \pm 0.32^\dagger$ | $-0.51 \pm 0.20$ | $-0.57 \pm 0.19$ |

LIME and SHAP produced the highest pairwise consistency scores in all four datasets, indicating stronger agreement between the two RF-based post-hoc attribution methods than between either method and TabNet. The strength of this agreement varied across datasets: it was highest for ESDRPD (0.66 ± 0.32), moderate for PDHS (0.46 ± 0.23), and weaker for Breast cancer (0.16 ± 0.42) and Diabetes retinopathy (0.08 ± 0.49).

Comparisons involving TabNet produced negative mean consistency scores across datasets. Because TabNet rankings are obtained from the native TabNet model, whereas LIME and SHAP explain the selected RF focal predictor, these values should be interpreted as cross-configuration rank divergence rather than disagreement among explainers applied to the same predictive function.

Overall, the consistency results suggest that LIME and SHAP provide the strongest observed feature-ranking alignment under the RF focal predictor, while TabNet yields a distinct ranking profile under its native model configuration.

##### Robustness

The robustness scores for LIME, SHAP, EBM, and TabNet are reported in Table 8. Results are summarized using mean ± standard deviation over test instances to capture both average sensitivity and variability of explanations under small input perturbations.

**Table 8.** Robustness of XAI method–predictor configurations measured as sensitivity of explanations to input perturbations, reported as mean ± standard deviation over test instances. LIME and SHAP explain the selected RF focal predictor, whereas EBM and TabNet are evaluated through native model-specific interpretability. Lowest values are marked descriptively with ^†^. Standard deviations are not shown when they round to 0.000.

| Method | PDHS<br>(C-section) | Diabetes<br>retinopathy | Breast<br>cancer | ESDRPD |
| --- | --- | --- | --- | --- |
| LIME | $0.0477 \pm 0.0338$ | $0.0428 \pm 0.0197$ | $0.0422 \pm 0.0202$ | $0.0672 \pm 0.0272$ |
| SHAP | $0.0002 \pm 0.0004$ | $0.0132 \pm 0.0219$ | $0.0078 \pm 0.0193$ | $0.0002 \pm 0.0009$ |
| EBM | $0.0000 \pm 0.0010^\dagger$ | $0.0021 \pm 0.0103$ | $0.0175 \pm 0.0388$ | $0.0000^\dagger$ |
| TabNet | $0.0006 \pm 0.0040$ | $0.0009 \pm 0.0106^\dagger$ | $0.0000 \pm 0.0007^\dagger$ | $0.0201 \pm 0.0343$ |

EBM and TabNet showed low sensitivity scores in their native model-specific explanation configurations under the fixed Gaussian-jitter protocol. EBM in particular exhibited near-zero mean sensitivity with negligible variance, indicating stable native explanations under this numerical perturbation setting. TabNet also produced low sensitivity scores in most datasets, although these results should be interpreted alongside its dataset-specific predictive performance, particularly for Breast cancer and ESDRPD (see Table 3).

For the RF focal predictor, SHAP generally showed low sensitivity, with moderate variability in Diabetes retinopathy and Breast cancer. In contrast, LIME was more sensitive to the perturbation and exhibited larger standard deviations, indicating greater instance-level instability in its local surrogate explanations.

Overall, the robustness results provide a comparative view of explanation sensitivity under a fixed numerical perturbation protocol. They should be interpreted as benchmark stability scores rather than as evidence of robustness under domain-valid clinical perturbations.

#### Evaluation of rule-based XAI method

##### Precision, coverage, simplicity, and robustness

The quantitative results for Anchors are reported in Table 9, summarizing how rule-based explanations vary with different precision thresholds. All metrics are reported as mean ± standard deviation over test instances to capture both average behavior and instance-level variability. In Anchors, the threshold controls the minimum acceptable precision for a rule to be extracted. While higher thresholds increase rule reliability (precision), they reduce the number of data points covered (coverage) and typically increase rule complexity (simplicity), as more conditions are needed to meet the stricter criteria.

**Table 9.** Precision, coverage, simplicity, and robustness of Anchors across datasets, reported as mean ± standard deviation over test instances. Standard deviations are not shown when they round to 0.000.

| Dataset | Threshold | Precision | Coverage | Simplicity | Robustness |
| --- | --- | --- | --- | --- | --- |
| PDHS<br>(C-section) | 0.90 | $0.96 \pm 0.04$ | $0.29 \pm 0.14$ | $2.35 \pm 4.05$ | $0.39 \pm 0.41$ |
| | 0.95 | $0.97 \pm 0.04$ | $0.24 \pm 0.17$ | $2.81 \pm 4.68$ | $0.40 \pm 0.41$ |
| | 0.99 | $0.99 \pm 0.04$ | $0.19 \pm 0.19$ | $3.65 \pm 5.13$ | $0.47 \pm 0.38$ |
| Diabetes<br>retinopathy | 0.90 | $0.91 \pm 0.04$ | $0.02 \pm 0.03$ | $11.01 \pm 9.38$ | $0.54 \pm 0.24$ |
| | 0.95 | $0.94 \pm 0.05$ | $0.01 \pm 0.01$ | $13.89 \pm 10.41$ | $0.56 \pm 0.20$ |
| | 0.99 | $0.96 \pm 0.06$ | $0.01 \pm 0.01$ | $16.89 \pm 10.79$ | $0.56 \pm 0.18$ |
| Breast<br>cancer | 0.90 | $0.99 \pm 0.02$ | $0.22 \pm 0.11$ | $2.03 \pm 0.17$ | $0.72 \pm 0.18$ |
| | 0.95 | $0.99 \pm 0.01$ | $0.22 \pm 0.09$ | $2.05 \pm 0.25$ | $0.72 \pm 0.21$ |
| | 0.99 | 1.00 | $0.18 \pm 0.08$ | $2.37 \pm 0.57$ | $0.70 \pm 0.22$ |
| ESDRPD | 0.90 | $0.93 \pm 0.03$ | $0.36 \pm 0.14$ | $2.12 \pm 1.19$ | $0.33 \pm 0.38$ |
| | 0.95 | $0.99 \pm 0.02$ | $0.21 \pm 0.10$ | $3.22 \pm 1.91$ | $0.45 \pm 0.34$ |
| | 0.99 | $1.00 \pm 0.01$ | $0.19 \pm 0.11$ | $3.60 \pm 2.04$ | $0.47 \pm 0.34$ |

Across all datasets, increasing the threshold improves rule precision, often reaching values of ≥ 0.99, but leads to a substantial reduction in coverage, in some cases explaining as little as 1% of instances (e.g., Diabetes retinopathy). This highlights that very precise rules typically apply only to a small subset of instances. Importantly, the relatively small standard deviations observed for precision indicate that Anchors reliably enforces the desired precision expectation level across instances once a threshold is fixed.

Coverage, in contrast, exhibits low mean values and modest variability across datasets, particularly for Diabetes retinopathy, where coverage remains consistently low across thresholds. This suggests that the narrow applicability of Anchors rules is not driven by a few outlier instances, but is a systematic consequence of enforcing high-precision constraints. In contrast, datasets such as PDHS and Breast cancer retain moderate coverage at lower thresholds, indicating that broader rules can be obtained when the underlying decision structure is simpler.

As thresholds increase, rule length generally increases, reducing compactness and requiring more feature conditions to satisfy stricter precision requirements. The reported standard deviations indicate substantial instance-level variability in rule length, particularly for datasets exhibiting higher heterogeneity, such as Diabetes retinopathy and PDHS, where simplicity standard deviations are comparable to or larger than the mean. This indicates that while some instances can be explained using relatively compact rules, others require substantially more conditions to achieve the same precision level, which has implications for human interpretability and practical usability.

Robustness generally remains moderate and, in some datasets, worsens slightly at stricter precision thresholds (lower values indicate higher robustness against noise), suggesting increased sensitivity of longer rules to small perturbations. The moderate standard deviations observed for robustness suggest that this sensitivity varies across instances, reflecting heterogeneity in how tightly rule conditions bind to individual patient profiles. Compared with the low sensitivity observed for native EBM explanations, Anchors rules showed greater perturbation sensitivity, particularly at higher thresholds.

Overall, these results reinforce a fundamental trade-off in rule-based explainability: *precision versus coverage versus simplicity versus robustness*. Anchors excels at producing human-readable, high-precision rules for narrowly defined subpopulations, but at the cost of limited coverage, variable rule complexity, and reduced robustness under noise. Compared to attribution-based methods such as LIME and SHAP, Anchors offers more actionable (human-readable) rule-based explanations, but for a smaller and more selective set of cases.

### Descriptive plausibility check of global feature profiles

#### Scope and interpretation

This subsection reports a descriptive plausibility check of global feature profiles. It is intended to contextualize model behavior by comparing aggregated explanation summaries with risk factors reported in prior literature. This is not a clinical validation analysis: no clinician-led review, external validation, causal assessment, or subgroup analysis was conducted. Accordingly, the results should be interpreted as descriptive face-validity evidence only, not as evidence of clinical utility, clinical significance, or medical decision validity.

To contextualize model behavior, we examined global feature-importance outputs from EBM, SHAP, and TabNet across the four healthcare tabular datasets. For the RF focal explainee, we also compared conventional RF global importances, permutation importance (PFI) and Gini/MDI, with the corresponding global explanation summaries as a descriptive cross-check of top-ranked signals (see Appendix, Figs. 3–10). Below, we summarize the main feature groups highlighted by each method and relate them cautiously to prior clinical literature.

#### PDHS (C-section) dataset

**EBM:** Key features include *Births in the last five years & Had Previous C-section*, *Had Previous C-section*, *Number of household members & Mothers BMI*, and *Age of Mother & Number of ANC Visits*.

**SHAP:** Important features highlighted are *Had Previous C-section*, *Suffered by Domestic Violence*, *Number of ANC Visits*, and *Type of place of residence*.

**TabNet:** Significant features identified are *Births in the last five years*, *Reading Newspaper or Magazine*, *Had Previous C-section*, and *Household Toilet Facility*.

**RF baseline (tree importances):** The RF summaries place *Had Previous C-section* among the dominant signals, with *Number of ANC Visits* and maternal-demographic factors also appearing prominently. This broadly overlaps with the EBM and SHAP summaries, while SHAP also assigns high importance to *Domestic Violence*, a socio-behavioural variable not always ranked at the top by RF split-based summaries (Appendix, RF: Fig. 3; SHAP/EBM/TabNet: Fig. 4).

These features are qualitatively consistent with factors reported in maternal health research. For instance, prior C-sections and antenatal care have been associated with surgical delivery outcomes, while domestic violence and household conditions reflect broader socioeconomic risks [25, 51, 52].

#### Diabetes retinopathy dataset

**EBM, SHAP, TabNet:** EBM, SHAP, and TabNet all highlight variants of *ma* (microaneurysms) and *ex* (exudates) features, such as *ma0.5*, *ex0.5*, and *exm1*, among the dominant signals.

**RF baseline (tree importances):** RF PFI and Gini/MDI also highlight lesion-related features, including *microaneurysms (ma*)* and *exudates (ex*)*, as important signals. This provides a descriptive cross-check against the

SHAP/EBM/TabNet summaries, which similarly emphasize retinal lesion features (Appendix, RF: Fig. 5; SHAP/EBM/TabNet: Fig. 6).

This pattern is consistent with prior literature on diabetes retinopathy, where microaneurysms and retinal exudates are reported as important diagnostic indicators [53, 54].

#### Breast cancer dataset

**EBM, SHAP, TabNet:** EBM, SHAP, and TabNet highlight geometric and morphological features, including *Mean/Worst concave points*, *Texture*, *Area*, and *Perimeter*.

**RF baseline (tree importances):** RF importances (PFI and Gini/MDI) surface geometric/morphological variables, *concave points (mean/worst)*, *area*, *perimeter*, and texture-related measures, consistent with EBM/SHAP rankings. This overlap with morphology-related variables provides descriptive face-validity for the global signals (Appendix, RF: Fig. 7; SHAP/EBM/TabNet: Fig. 8).

These features are consistent with breast cancer literature on malignancy-related morphology, including concavity, area, perimeter, and texture measures [55, 56].

#### ESDRPD dataset

**EBM, SHAP, TabNet:** EBM, SHAP, and TabNet highlight symptom-related features such as *Polyuria*, *Polydipsia*, *Gender*, and *Partial paresis*, with TabNet additionally emphasizing features such as *Muscle stiffness* and *Sudden weight loss*.

**RF baseline (tree importances):** RF PFI and Gini/MDI also rank classic symptom variables, including *Polyuria*, *Polydipsia*, *Gender*, and *Partial paresis*, among the leading contributors, with occasional emphasis on related neurological or metabolic features.

This provides a descriptive cross-check against the symptom profile highlighted by the global explanation summaries (Appendix, RF: Fig. 9; SHAP/EBM/TabNet: Fig. 10).

These symptoms are consistent with prior literature on early diabetes indicators and diabetes-related complications [57–59].

Overall, the global explanation summaries from EBM, SHAP, TabNet, and RF importances provide descriptive numerical context for model behavior. In several cases, different methods highlight broadly similar feature groups; in others, they emphasize distinct signals. Such convergence and divergence should be interpreted as exploratory evidence for further investigation, not as validated clinical findings. The qualitative consistency with previously reported risk factors supports a face-validity or plausibility interpretation, but does not establish clinical validity, causal relevance, subgroup generalisability, or usefulness in medical decision-making. Figures supporting these descriptive analyses are presented in the Appendix (see Figs. 3–10).

## Discussion

This study presents a comparative, metric-based evaluation of five explainable AI (XAI) methods, SHAP, LIME, Anchors, EBM, and TabNet, on structured healthcare datasets. Using six evaluation metrics, fidelity, simplicity, consistency, robustness, precision, and coverage, we assessed explanation behavior under the method–metric applicability design described in Table 2. Post-hoc results for LIME, SHAP, and Anchors are conditional on the selected RF focal predictor, whereas EBM and TabNet results correspond to their own native predictors. In addition, we juxtaposed conventional Random Forest (RF) global feature importances, permutation (PFI) and Gini/MDI, as contextual baselines against SHAP/EBM/TabNet global summaries.

### Method-level interpretations and trade-offs

#### Fidelity

SHAP obtained score fidelity of 1.000 across all datasets (Table 5). This reflects the expected exact additive reconstruction property of TreeSHAP for tree ensembles, rather than an empirical superiority finding over surrogate-based methods. In contrast, LIME’s fidelity reflects the approximation quality of its local surrogate model, which can vary across instances and datasets.

#### Simplicity

The simplicity results describe explanation compactness across method–predictor configurations rather than a strict ranking of methods under a common predictive function. TabNet produced compact native explanations in several settings, particularly at lower thresholds (0.01 and 0.05), but these results should be interpreted alongside its dataset-specific predictive performance. LIME produced compact RF-based local explanations in some settings and often saturated at a fixed number of selected features once the threshold constraint became binding. SHAP explanations were generally denser, while EBM produced more feature-rich native explanations (Table 6). Among rule-based explanations, Anchors produced short rules in several datasets (Table 9), but rule length varied substantially across instances and coverage remained limited at higher precision thresholds.

#### Consistency

LIME and SHAP showed the highest pairwise rank agreement across datasets, although the magnitude of agreement was dataset-dependent. Agreement was strongest for ESDRPD and PDHS, but weaker for Breast cancer and Diabetes retinopathy (Table 7). Comparisons involving TabNet were consistently negative, reflecting divergence between RF-based post-hoc rankings and native TabNet rankings.

These findings suggest that inter-explainer consistency is useful as a diagnostic measure of rank alignment, but should not be interpreted as evidence of explanation correctness.

#### Robustness

Robustness scores should be interpreted as numerical sensitivity measures under the fixed perturbation protocol and in relation to the underlying predictor. EBM and TabNet showed low perturbation sensitivity in their native explanation configurations, whereas SHAP and LIME were evaluated on the selected RF focal predictor (Table 8). For RF-based explanations, SHAP generally showed lower sensitivity than LIME, while LIME exhibited larger variability across instances.

Anchors also showed non-trivial sensitivity under perturbation (Table 9), with robustness varying across datasets and thresholds. These results support the usefulness of robustness as a comparative stress test for explanation stability, but not as a model-independent or clinically validated ranking of explanation methods.

Fig 2 demonstrates this instability using two LIME explanations for the same ESDRPD instance, once with the original input and once with a minor perturbation. While the predicted class remains the same, the explanations shift significantly in content and magnitude. This perturbation was applied by adding small Gaussian noise (N (0, 0.01^2^)) to the original input instance, as described in the *Materials and methods* section under Robustness. The example therefore illustrates local explanation instability under the benchmark perturbation protocol, rather than instability under clinically realistic patient change.

**Fig 2.**
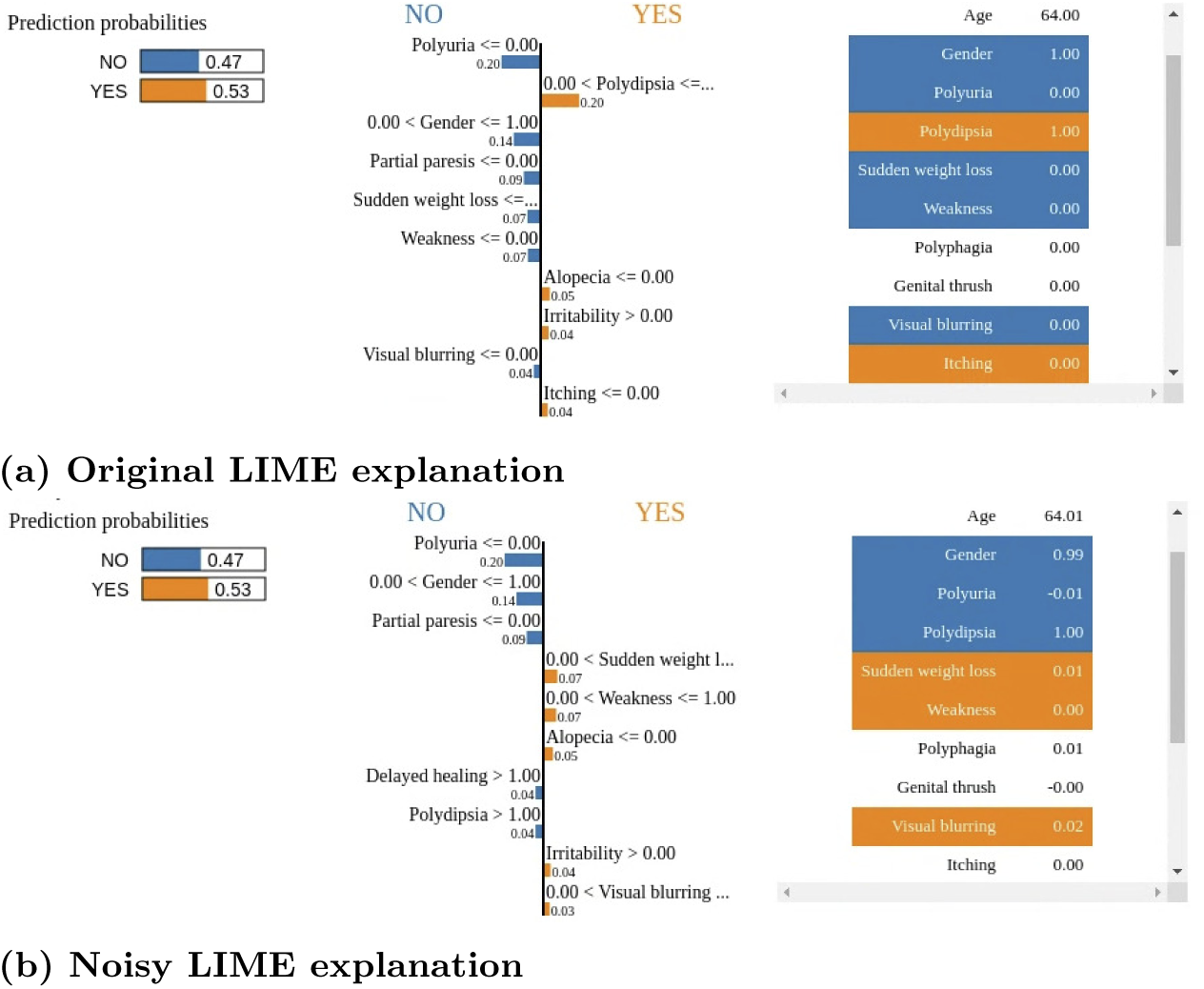
Original and perturbed/noisy LIME explanations for an ESDRPD instance. The perturbation is element-wise Gaussian jitter, *ɛ* ∼ N (0, 0.01^2^), applied to the numerically encoded input representation with no model refitting.

In parallel, Anchors generated different rule sets for the same instance under noise:

- **Original Anchor Rule:** *Age* ≤ 47.50 AND *Polyuria* ≤ 0.00 AND *V isual blurring* ≤ 0.00 AND *Gender >* 0.00 AND *Itching >* 0.00
- **Noisy Anchor Rule:** *Polydipsia >* 0.00 AND *Irritability >* 0.00 AND *Obesity* ≤ 0.00 AND *Age >* 58.00

These results show that post-hoc explainers relying on local sampling (LIME) or greedy rule induction (Anchors) can be fragile under perturbation, especially in datasets with closely correlated features or small decision boundaries.

##### Precision and coverage

Anchors excelled in producing precise rules with interpretable conditions, making it valuable in applications like subgroup-style model auditing. However, the cost of this precision is low coverage, limiting its utility for broad model auditing (Table 9). Reporting mean ± standard deviation clarifies that these behaviors are systematic rather than driven by a few outliers: precision shows relatively small variability once a threshold is fixed, whereas coverage is consistently low (and only modestly variable), especially in Diabetes retinopathy. Rule complexity (simplicity) exhibits large instance-level dispersion in some datasets (e.g., PDHS and Diabetes retinopathy), indicating that some instances can be captured by compact rules while others require substantially longer rules to meet the same precision threshold. Robustness also shows moderate variability, suggesting heterogeneous stability under perturbations across instances. This trade-off should be considered when using Anchors for local model auditing or subgroup-style interpretation in healthcare tabular settings.

##### Triangulation with traditional tree importances

Across datasets, RF PFI and Gini/MDI broadly corroborate the global signals surfaced by SHAP/EBM/TabNet: (i) *PDHS (C-section)*, strong emphasis on *Had Previous C-section* with *ANC visits*/maternal-demographic factors also high; SHAP further lifts socio-behavioral risk (*Domestic Violence*). (ii) *Diabetes retinopathy*, lesion markers (*ma\**, *ex\**) dominate across RF and SHAP/EBM. (iii) *Breast cancer*, geometric/morphological variables (*concave points*, *area*, *perimeter*, texture) agree across methods. (iv) *ESDRPD*, classic symptoms (*Polyuria*, *Polydipsia*, *Gender*, *Partial paresis*) align. Where differences arise, they are typically in the ordering of secondary factors rather than complete disagreement, and may reflect known properties of impurity-based scores, such as bias toward high-cardinality splits, or permutation effects under correlated features. This descriptive cross-check contextualizes the global patterns while reinforcing the need for local metrics such as fidelity and robustness.

##### Clinical relevance and scope of global explanations

The global explanation summaries discussed above (e.g., aggregated SHAP importances, EBM main-effect functions, and TabNet attention profiles) are intended as *descriptive, numerical context* rather than as statistically grounded feature selection, causal analysis, or clinical validation. Their role is to summarize model behavior and to support a qualitative alignment check against well-established risk factors reported in the clinical literature. No clinician-led qualitative review was conducted, and we do not claim clinical significance or medical decision validity based on these results alone.

Clinician-in-the-loop validation is outside the scope of this methodological study and is identified as a direction for future work. These summaries should therefore be interpreted as interpretability-oriented diagnostics that complement, rather than replace, domain-expert evaluation.

### Best practices and practical recommendations

Based on our evaluation, we recommend the following XAI method selection guidelines tailored to typical healthcare scenarios. These recommendations are intended as practical heuristics for model debugging and interpretability workflows, rather than as guidance for clinical deployment.

- **For fidelity-critical model-auditing workflows:** SHAP is appropriate for tree-based models when exact or near-exact local additive reconstruction is required; in this study, this was demonstrated for the RF focal predictor (see Table 5). For example, in oncology risk stratification tools or ICU early warning systems, precise probability estimates are essential for triggering critical interventions like escalation of care. Misalignment between explanations and model outputs may result in incorrect risk thresholds.
- **For simplicity and rapid user-facing explanations:** Consider compact explanation configurations such as LIME, Anchors, or native TabNet explanations (see Tables 6 and 9), provided that the underlying predictive model first meets the required performance threshold for the task. These can be suitable in resource-constrained settings, such as mobile-based screening tools for frontline triage systems, where clinicians or health workers need fast, intuitive reasoning to communicate decisions effectively to patients.
- **For low sensitivity under the benchmark perturbation protocol:** Consider EBM or TabNet when their predictive performance is adequate for the target dataset, as these models produced low sensitivity scores in their native explanation configurations under the fixed Gaussian-jitter protocol (see Table 8). These findings are most useful for model debugging and comparative sensitivity analysis; domain-calibrated perturbations would be needed before drawing conclusions about stability under clinically plausible variation.
- **For local, rule-based decision support:** Apply Anchors, noting its narrow applicability. Anchors is effective for tasks requiring precise rule extraction (see Table 9), such as auditing medical eligibility criteria, drug contraindication alerts, or subgroup-specific recommendations where interpretability outweighs global applicability.
- **When using tree ensembles, sanity-check with RF importances first, then escalate to SHAP when stakes are high:** PFI/MDI provide a fast global ranking for model debugging, but can misorder correlated or high-cardinality features; SHAP (TreeSHAP) offers exact additive local reconstruction for compatible tree models and more reliable global aggregation when decisions hinge on calibrated probabilities or safety-critical thresholds.

These recommendations are not limited to healthcare. The evaluation framework and insights can be applied to domains such as finance, insurance, fraud detection, marketing, and industrial systems, where tabular data is prevalent, and explainability is critical for regulatory compliance, operational trust, or model debugging.

### Descriptive alignment with clinical literature

Across datasets, SHAP, EBM, and TabNet surfaced global features that were qualitatively consistent with risk factors reported in prior clinical studies, including maternal indicators for C-sections (Fig. 4), retinal lesion markers for diabetes retinopathy (Fig. 6), tumor morphology measures in breast cancer (Fig. 8), and early diabetes symptoms in ESDRPD (Fig. 10). These alignments are detailed in the *Results* section and should be interpreted as a descriptive plausibility check that contextualizes model behavior, rather than as feature validation, causal evidence, clinical significance, or clinical utility.

Conversely, lower-fidelity or less robust explainers (e.g., LIME and Anchors under perturbation) can produce explanations that vary substantially across instances, underscoring the value of metric-guided explainer selection and sanity checks in high-stakes tabular settings.

### Limitations and future work

This study focuses on publicly available, tabular healthcare datasets and does not extend to imaging, time-series, or longitudinal modalities that are increasingly prominent in clinical AI. We evaluate six quantitative metrics; however, complementary qualitative dimensions lie outside our scope and warrant future user studies.

#### Human-centred evaluation

We assess explanation quality via quantitative, reproducible proxies, including fidelity, simplicity, inter-explainer consistency, robustness, and precision–coverage evaluated across datasets. Direct human-centred evaluation (e.g., clinician task accuracy, completion time, error rate, SUS/NASA–TLX, perceived trust or appropriateness) is beyond the scope of this study and planned as future work with domain experts. Direct clinician involvement in reviewing explanation outputs and assessing medical plausibility was not part of this study and is identified as a necessary direction for future work.

#### Scope of coverage

Our emphasis is on a principled *evaluation framework* and transparent operationalization of quantitative XAI metrics, not exhaustiveness over explainer families. Metrics are reported only where theoretically appropriate to each explainer’s output type (e.g., rank-based consistency for per-instance attribution vectors; precision/coverage for rule sets). This policy avoids cross-paradigm conflation and supports reuse; future extensions can broaden method families under the same protocol.

#### Design choices

Our post-hoc comparison is performed on a *single focal predictor* selected after benchmarking via a pre-specified average-rank rule (F1 primary; accuracy tie-breaker). This design controls model-induced variance so that differences among LIME, SHAP, and Anchors are attributable to the explainer rather than to changes in the underlying decision function. The selected RF model is not the best-performing classifier on every dataset, including Diabetes retinopathy; therefore, the post-hoc findings should be interpreted as conditional on this focal explainee rather than as model-invariant explainer behavior. Replicating the protocol across alternative top-ranked predictors such as XGB or GBC would be valuable for studying explainer–model interactions, but introduces an additional experimental axis beyond the present study boundary. EBM and TabNet are assessed via their inherent interpretability and should therefore be understood as native interpretable model configurations, not as explanations of the RF decision function. We also avoid extensive per-dataset hyperparameter tuning (fixed seeds; library defaults) to reduce confounding variability in explanation metrics. While this may affect absolute predictive scores, it improves comparability and reproducibility of explainer results.

#### Global tree importances

PFI and Gini/MDI are global summaries: they aid triangulation but neither guarantee local agreement with the model nor immunity to known biases (e.g., impurity-based bias toward high-cardinality features; permutation underestimation/instability with collinear predictors or leakage). Their sensitivity to data distribution and feature correlation further motivates our use of local, instance-level metrics (fidelity, robustness) for post-hoc explainers.

#### Perturbation model (robustness)

Robustness was quantified using small, zero-mean, element-wise Gaussian perturbations as a method-agnostic numerical sensitivity probe. This design supports controlled comparison across explainers and datasets, but it does not model clinically plausible or physiologically meaningful patient variation. Because the perturbation is applied to numerically encoded inputs, it does not preserve categorical levels, ordinal constraints, feature codebooks, or valid biomarker ranges. The robustness scores should therefore be interpreted as explanation sensitivity under a fixed computational stress test. Future work should complement this generic perturbation scheme with domain-calibrated perturbations that preserve categorical levels, respect feature ranges, and use biomarker- or codebook-aware variation.

#### Method breadth

We study widely used representatives, post-hoc feature-attribution methods (SHAP, LIME), a rule-based local explainer (Anchors), and inherently interpretable models (EBM, TabNet), rather than an exhaustive catalog. Other rule-list/decision-set learners and glass-box variants could be included; our open code is structured to accommodate such extensions.

#### Metric scope

The reported metrics are computed deterministically on a fixed test set, and the study is designed for descriptive, instance-level comparison rather than population inference. Accordingly, we report dispersion via standard deviation to characterize variability and potential failure modes across patients.

We consolidate and operationalize commonly cited quantitative metrics (fidelity, simplicity, consistency, robustness, and precision/coverage for rule-based explanations). These do not exhaust all meaningful criteria (e.g., human-centered measures such as usefulness or actionability), and some metrics are family-specific by design (e.g., precision/coverage for rules; rank-based consistency for single-feature attribution lists). Where relevant, partial sensitivity is reported directly in the results (e.g., multiple simplicity thresholds and Anchor precision levels), while broader sensitivity analyses are deferred to future work.

#### Future work

Future research should: (i) extend the benchmark to temporal and multimodal clinical datasets; (ii) run clinician-in-the-loop studies to assess perceived usefulness and cognitive effort; (iii) integrate additional explainer families, especially counterfactual and example-based methods, with appropriate, family-specific metrics (e.g., plausibility, feasibility, actionability); (iv) replicate the post-hoc analysis on alternative top-ranked predictors (e.g., boosted trees or a neural tabular model) to probe explainer–model interactions and assess cross-model generalizability; (v) harmonize evaluation between inherent and post-hoc explanations (e.g., restricting EBM consistency to main-effect functions or developing interaction-aware consistency measures); (vi) systematically characterize RF–SHAP divergence under controlled feature correlation and cardinality settings to identify regimes where traditional tree importances deviate most from TreeSHAP-based local additive attributions; and (vii) develop domain-calibrated perturbation schemes for robustness (biomarker- and codebook-aware ranges) and compare them to generic Gaussian jitter to assess clinical impact.

## Conclusion

We presented a reproducible, metric-driven *evaluation framework* for explainable AI (XAI) on clinical tabular models, focusing on the standardized operationalization and comparative application of established quantitative metrics. Rather than proposing a new explainer or evaluation metric, we formalized six established quantitative metrics, fidelity, simplicity, consistency, robustness, precision, and coverage, into explicit equations, mapped their applicability across method families, and released open-source code with a pre-specified focal-model selection protocol to reduce confounds. Applying the framework to five widely used approaches (LIME, SHAP, Anchors, EBM, TabNet) over four healthcare datasets yields consistent patterns under the stated focal-model protocol: SHAP (TreeSHAP) provides exact score fidelity for tree ensembles and near-perfect decision fidelity; LIME produces sparser (simpler) local rationales but with lower fidelity and robustness; TabNet, when assessed through its native feature-selection mechanism, often yields the fewest dominant features across thresholds, although this must be interpreted alongside its dataset-specific predictive performance; EBM and TabNet show low perturbation sensitivity in their native model configurations; and Anchors supplies high-precision, human-readable rules that exhibit the canonical precision–coverage trade-off as thresholds tighten. Reporting mean ± standard deviation reveals substantial instance-level variability for some explainers and metrics (e.g., LIME decision fidelity and TabNet/SHAP explanation sizes), highlighting that close mean values should be interpreted alongside dispersion rather than in isolation, particularly in the absence of inferential testing where stability and failure modes are the primary concern. LIME and SHAP show the strongest RF-based top-feature rank agreement among the evaluated pairs, although agreement varies across datasets, while global importance from EBM/SHAP/TabNet is qualitatively consistent with known risk factors as a descriptive plausibility check.

### Practical guidance

Match the explanation configuration to the evaluation priority: (i) *Fidelity-critical model auditing*: SHAP with compatible tree ensembles such as RF; (ii) *Compact local explanations*: LIME or Anchors, depending on whether feature attributions or rules are preferred; (iii) *Low sensitivity under small perturbations*: native EBM or TabNet explanations, provided predictive performance is adequate; and (iv) *High-precision rules for subgroups*: Anchors, accepting reduced coverage at higher precision thresholds.

### Scope and next steps

The framework targets tabular healthcare and is readily transferable to other high-stakes tabular domains. It does not claim exhaustiveness of methods or metrics; rather, it offers a transparent baseline that can be extended. Future work includes adding temporal and multimodal datasets, clinician-in-the-loop evaluations of usefulness and cognitive load, integrating counterfactual and example-based families with appropriate metrics, and probing explainer–model interactions by replicating the post-hoc analysis on alternative top-ranked predictors. Future robustness analysis can complement generic Gaussian jitter with domain-calibrated perturbations that preserve categorical levels, respect valid codebooks, and reflect clinically plausible biomarker ranges.

Finally, when tree ensembles are used, traditional global RF importances (Gini/MDI and permutation) are useful *descriptive cross-checks* but not substitutes for local fidelity-based evaluation; in our experiments, they qualitatively aligned with SHAP/EBM global patterns while SHAP additionally surfaced socio-behavioral risk factors that split-based importances may down-weight (see Appendix).

Overall, the framework turns narrative guidance into testable, quantitative practice, enabling controlled comparisons under explicit method–metric and focal-model assumptions and more defensible XAI selection in regulated settings.

## Author Contributions

**Conceptualization:** M. Atif Qureshi, Abdul Aziz Noor, Wael Rashwan, Arjumand Younus. **Data curation:** Abdul Aziz Noor, M. Atif Qureshi, Awais Manzoor. **Methodology – XAI Methods:** M. Atif Qureshi, Abdul Aziz Noor. **Methodology – XAI Metrics:** M. Atif Qureshi, Abdul Aziz Noor, Muhammad Deedahwar Mazhar Qureshi. **Software:** M. Atif Qureshi, Abdul Aziz Noor. **Formal analysis:** M. Atif Qureshi. **Investigation:** M. Atif Qureshi, Abdul Aziz Noor. **Supervision/Mentor of Student Authors:** M. Atif Qureshi, Wael Rashwan, Arjumand Younus. **Validation:** M. Atif Qureshi, Muhammad Deedahwar Mazhar Qureshi, Wael Rashwan, Arjumand Younus. **Visualization:** M. Atif Qureshi, Abdul Aziz Noor. **Writing – original draft:** M. Atif Qureshi, Abdul Aziz Noor. **Writing – review & editing:** M. Atif Qureshi, Muhammad Deedahwar Mazhar Qureshi, Awais Manzoor, Wael Rashwan, Arjumand Younus.

## Data and Code Availability

All code for metric definitions and experiments is available at https://github.com/matifq/XAI_Tab_Health. All datasets used are publicly available as cited in the manuscript.

## Data Availability

This study used publicly available, anonymized healthcare datasets, ensuring ethical compliance and data privacy. No new data was collected, and all analyses prioritized research integrity and reproducibility.

https://github.com/matifq/XAI_Tab_Health

## Appendix

**Fig 3.**
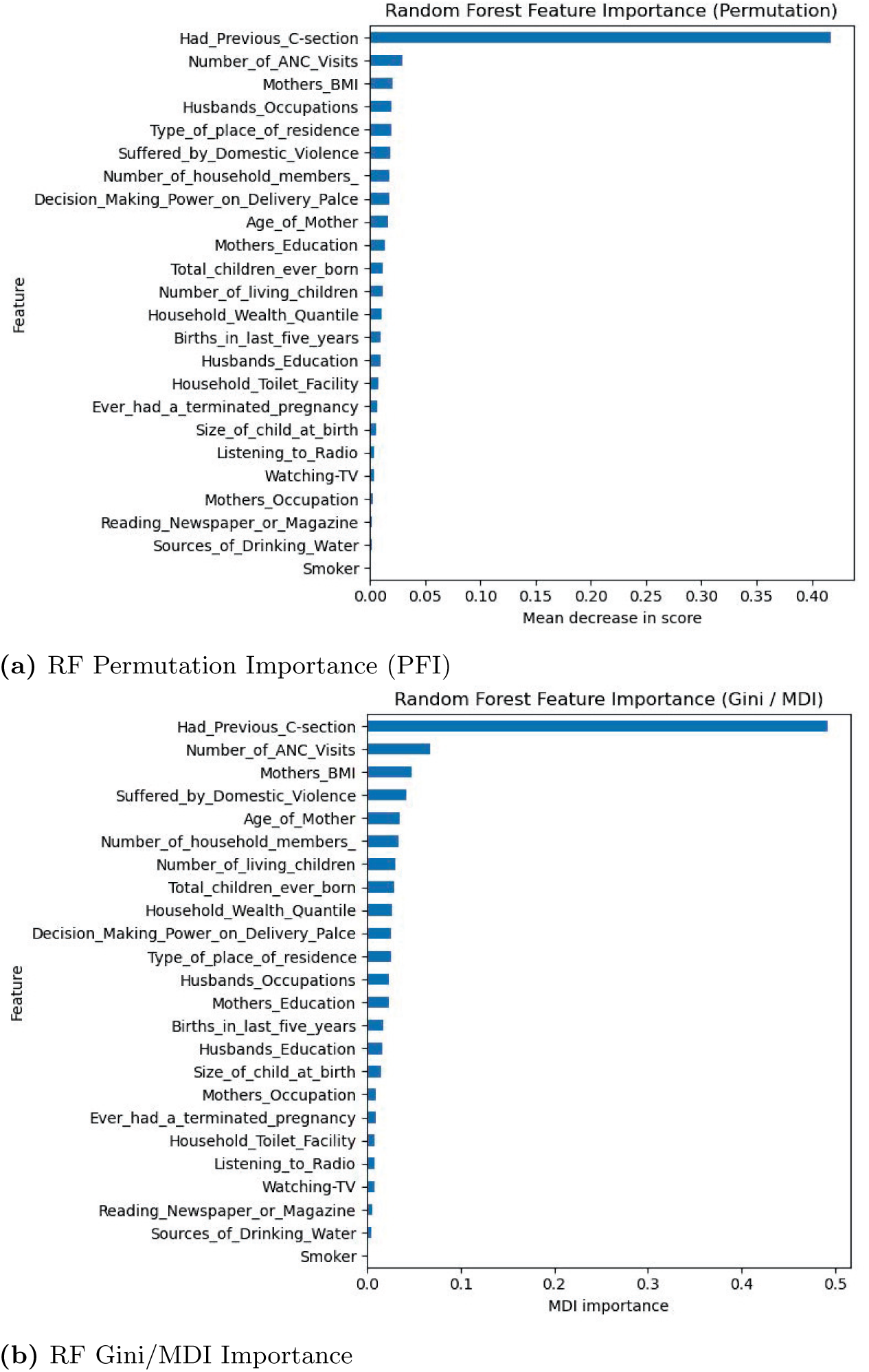
PDHS (C-section): RF global explanations.

**Fig 4.**
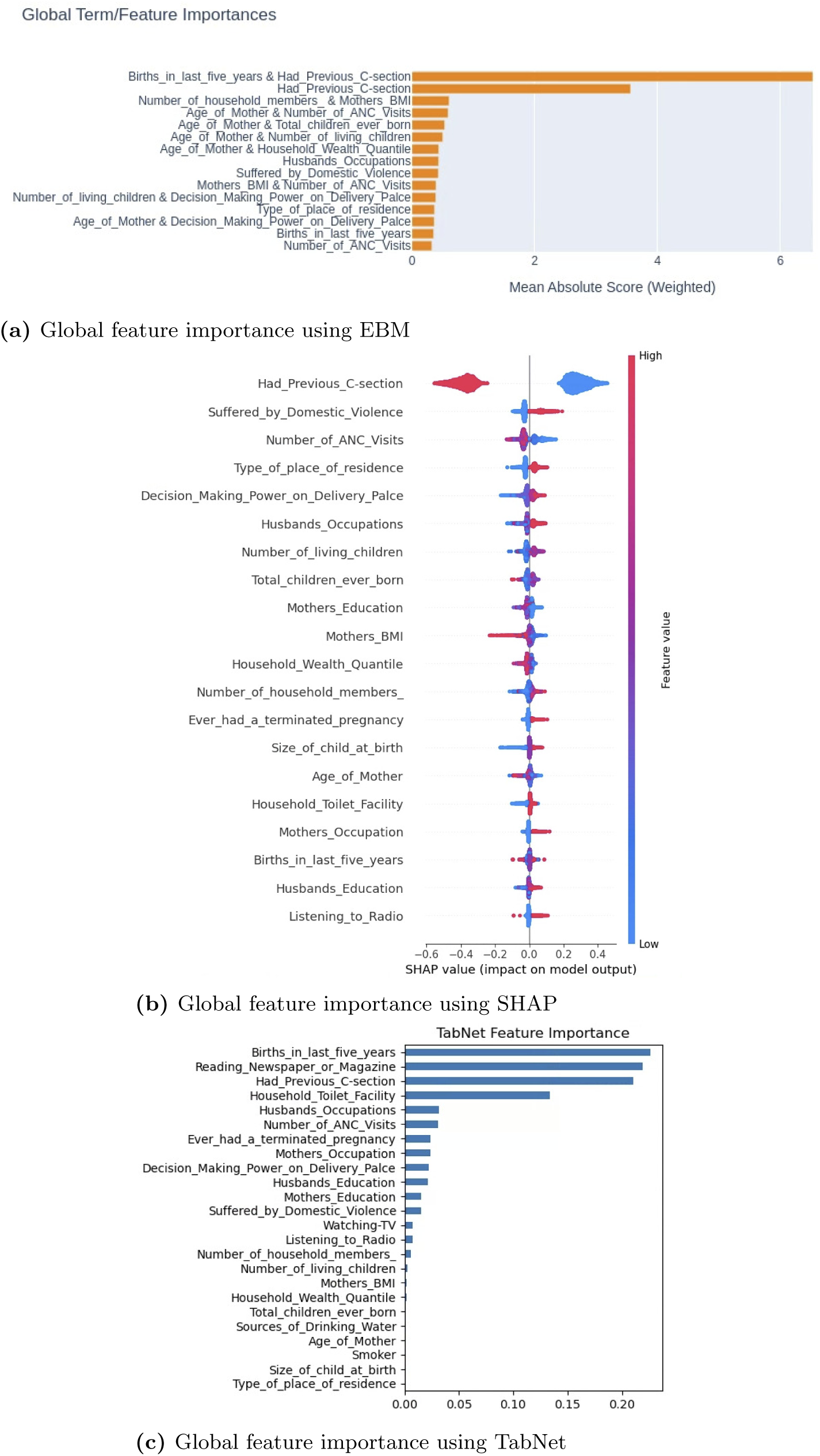
Global feature importance for PDHS (C-section) dataset using EBM, SHAP, and TabNet.

**Fig 5.**
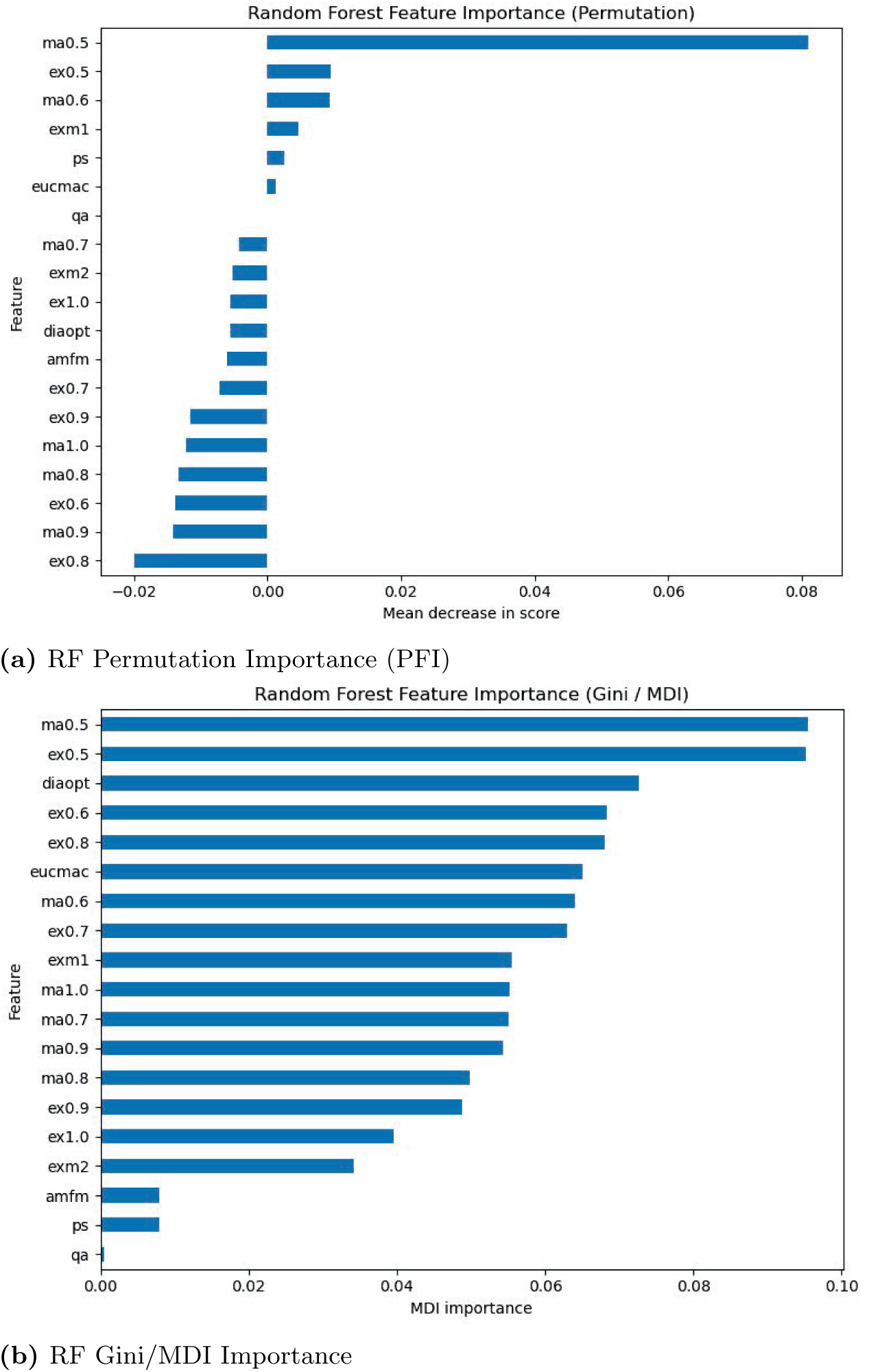
Diabetes retinopathy: RF global explanations.

**Fig 6.**
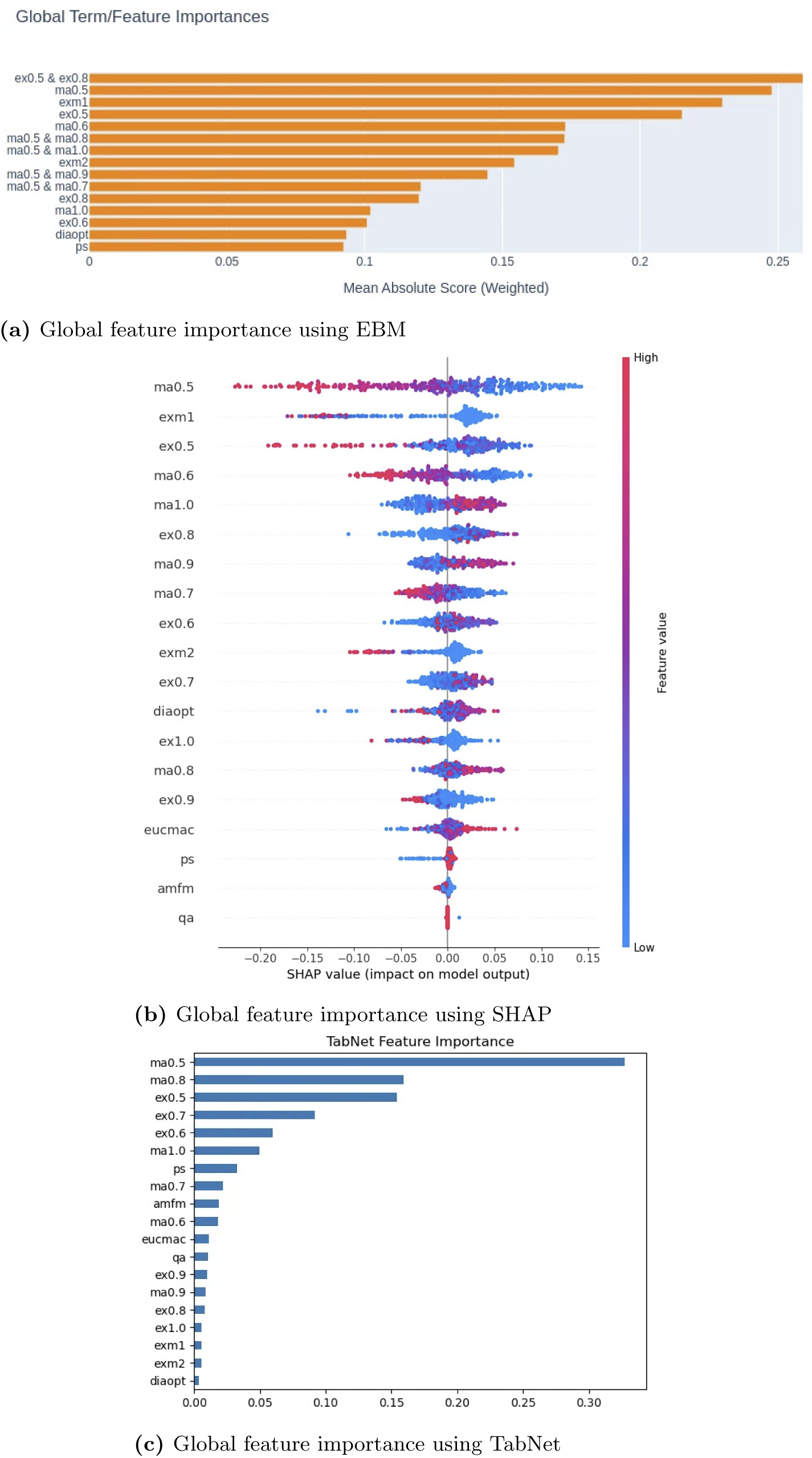
Global feature importance analysis for diabetes retinopathy dataset using EBM, SHAP, and TabNet.

**Fig 7.**
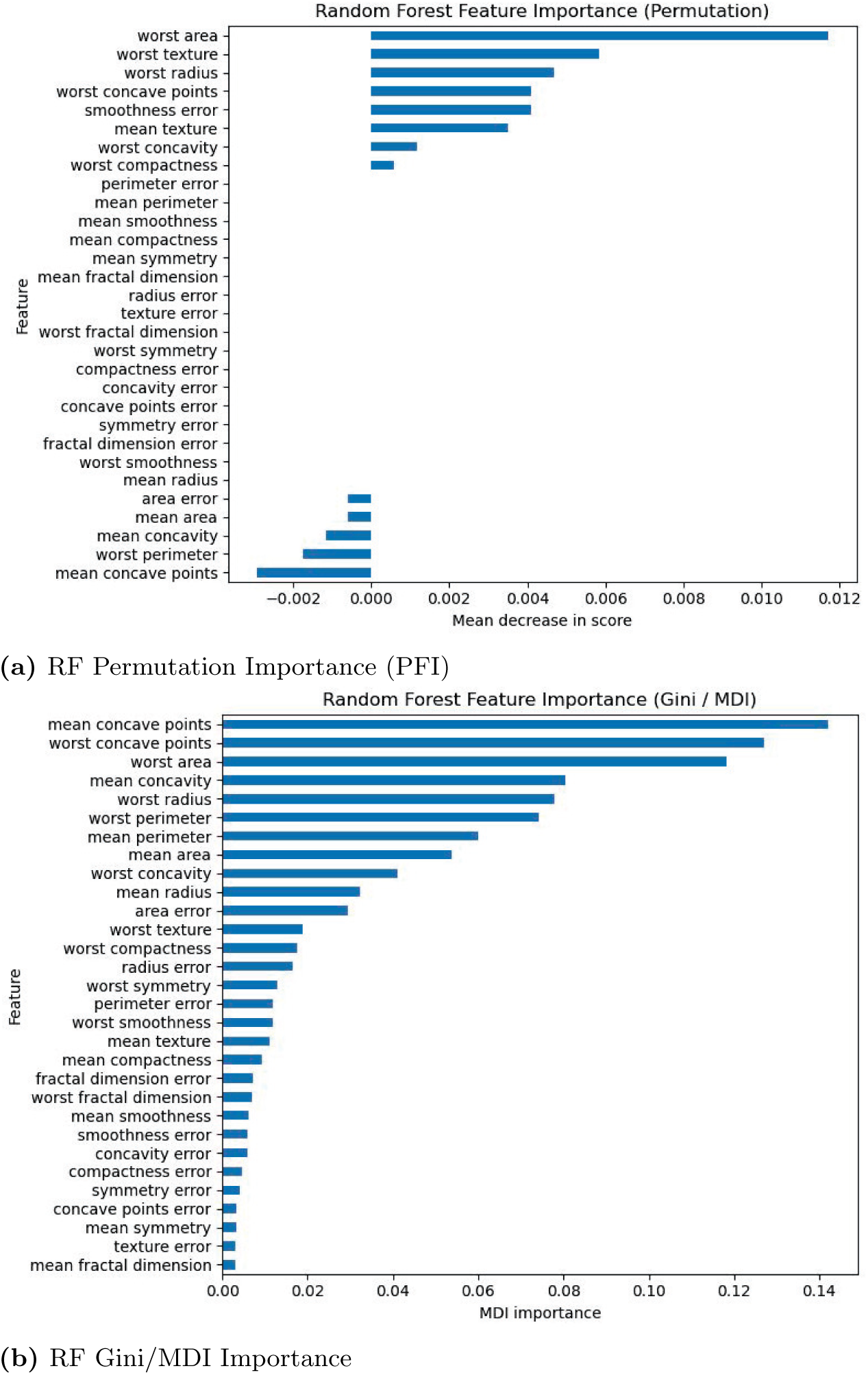
Breast cancer: RF global explanations.

**Fig 8.**
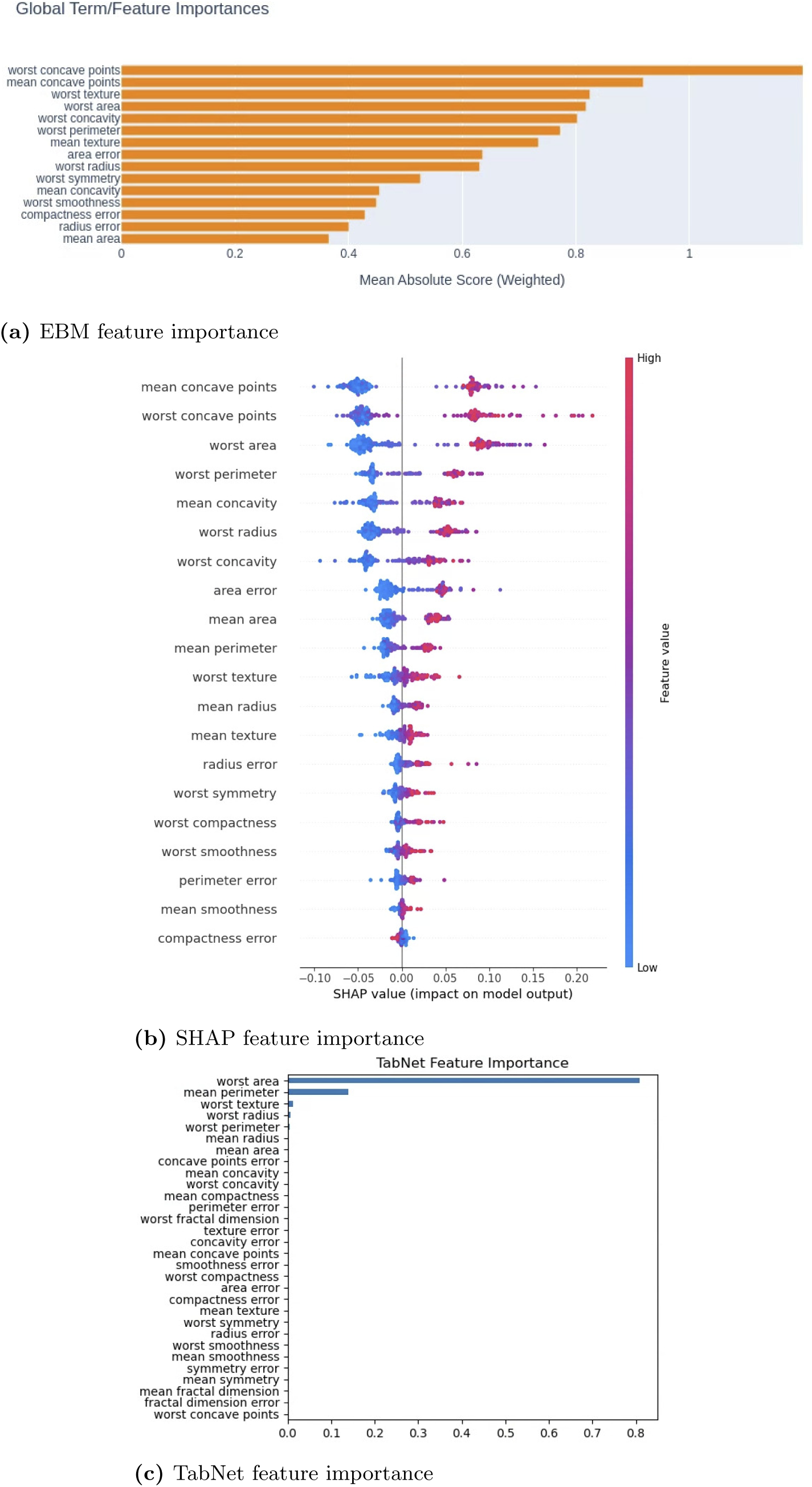
Global feature importance analysis for breast cancer dataset using EBM, SHAP, and TabNet.

**Fig 9.**
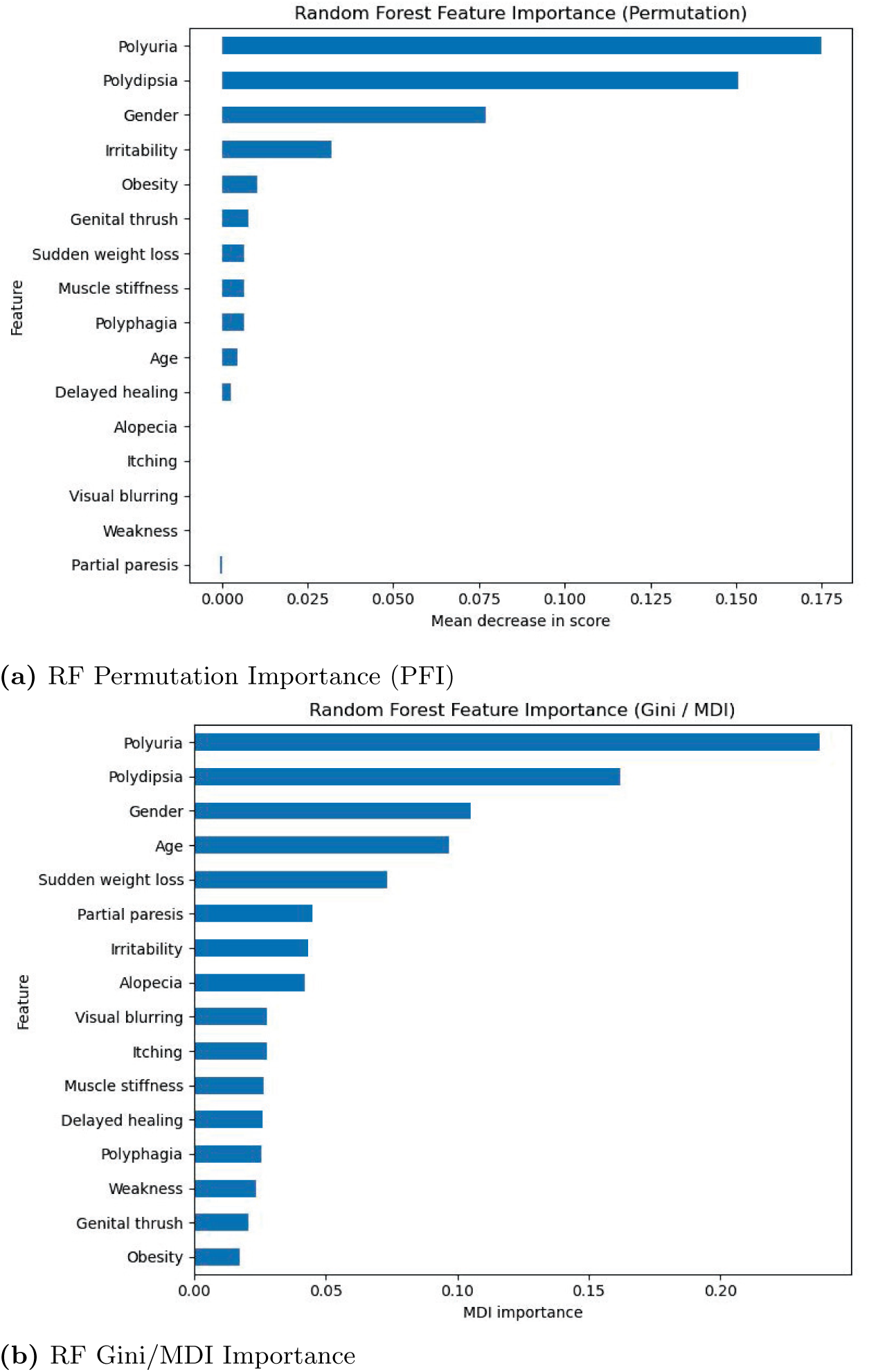
ESDRPD: RF global explanations.

**Fig 10.**
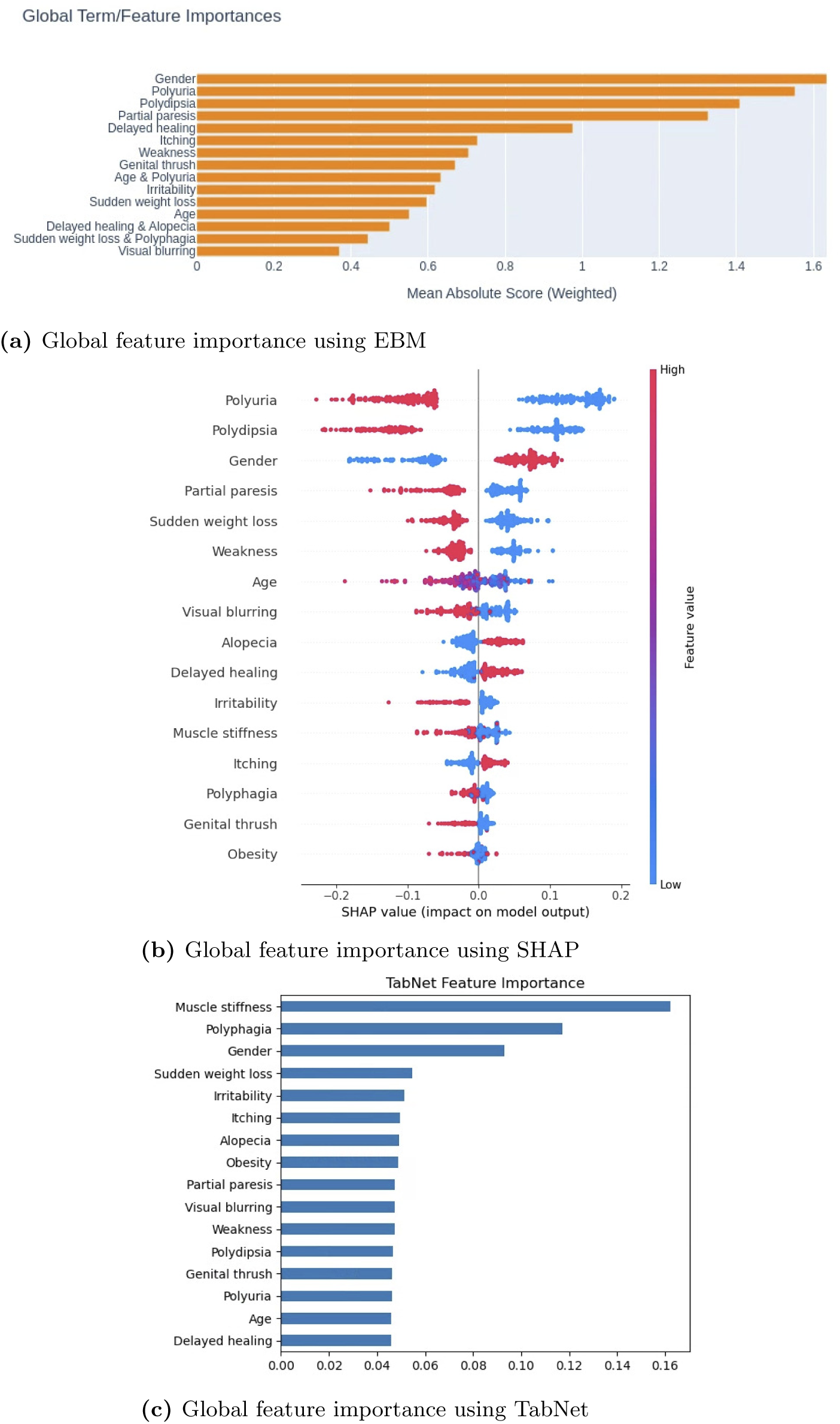
Global feature importance analysis for ESDRPD dataset using EBM, SHAP, and TabNet.

## Footnotes

1 or explainability

2 We report SHAP both locally (per-instance) and as aggregated global importances (mean absolute SHAP values), whereas LIME is analyzed as a local explainer only. Anchors is local and rule-based. EBM and TabNet are evaluated via their inherent (ante-hoc) interpretability; TabNet also exposes per-sample feature masks (local), with optional aggregation to summarize global trends.

3 TreeSHAP for tree ensembles; additive reconstruction is exact [34]. For non-tree models, KernelSHAP yields an additive approximation; the same fidelity equations apply, but we do not evaluate that variant here.

4 EBM (a GA^2^M) provides per-feature shape functions and optional *pairwise* interactions [42]; our rank-based consistency metric is defined for single-feature lists. To avoid conflating main effects with interactions [43], we do not compute consistency for EBM, leaving interaction-aware consistency for future work.

5 We use the *ℓ*_1_ norm; results are qualitatively similar with *ℓ*_2_. We do not divide by *|F |* so robustness reflects the *total* attribution shift per instance.

6 E.g., “*Age >* 45 AND *BMI >* 30” has length 2; adding “*Smoker* = *Y es*” gives length 3.

7 We use Jaccard dissimilarity on rule feature sets in Eq. (9); other set distances (e.g., Hamming on binary literal vectors) produce qualitatively similar trends.

8 Settings used in all experiments: random state= 42 for all models; RF: n estimators= 100, n jobs= *−*1; GBC: scikit-learn defaults; XGB: library defaults; EBM: library defaults; TabNet: seed= 42, verbose= 0. Full configuration files and code are available.

9 i.e., practitioners assess which threshold appears most suitable based on the dataset

